# Autoimmune interactions between the HLA-DQβ1_57_ polymorphism, T cell receptors, and microbial mimics of insulin in type 1 diabetes

**DOI:** 10.1101/2022.05.11.22274678

**Authors:** Arcadio Rubio García, Athina Paterou, Rebecca D. Powell Doherty, Laurie G. Landry, Mercede Lee, Amanda M. Anderson, Claire L. Scudder, Hubert Slawinski, Ricardo C. Ferreira, Dominik Trzupek, Agnieszka Szypowska, Luc Teyton, Nicola Ternette, Maki Nakayama, Linda S. Wicker, John A. Todd, Marcin L. Pekalski

## Abstract

Insulin autoimmunity and pancreatic islet β-cell destruction, mediated by T cells, underlie the pathogenesis of type 1 diabetes (T1D). However, the mechanisms by which genetic and environmental risk factors interact to cause T1D remain incompletely understood. Here, we show how variation in the human leukocyte antigen (HLA) class II region, notably DQβ1_57_, the strongest T1D genetic risk factor, induces a positive thymic selection bias that favors amino acid motifs with negatively charged side chains in the peptide-binding CDR3β region of CD4^+^ T cell receptors (TCRs). This bias was enriched in TCRs from T1D patients and predicted the proportion of anti-insulin infiltrates in pancreatic islets. We link host genetics and immunity to the environment by identifying over 100 gut bacterial proteins with significantly similar sequences to a primary epitope in T1D, insulin B_9–25_ peptide. We show that CD4^+^ T cells isolated from the pancreas of a newly diagnosed child with T1D recognised both insulin and a bacterial mimotope peptide, restricted by the most predisposing DQ molecule. Our results point to the evolution of certain microbial antigens towards mimicry in the regulation of immune tolerance to insulin.

## Introduction

Type 1 diabetes (T1D) is a common autoimmune disease characterized by a T cell-mediated destruction of pancreatic islet β cells, resulting in a lifelong reliance on exogenous insulin for metabolic control and survival. ^1^ Autoantibodies against insulin often emerge as the first pre-clinical sign of the disease, peaking in incidence between 9 and 18 months of age. ^2,3^ Before this period, the immune system and the pancreas undergo major developmental changes, regulated by host genetic variation and shaped by successive waves of mucosal microbial colonization. This process is particularly relevant in the gut, which harbors the greatest number of T cells in the body and shares lymph nodes with the pancreas, leading to immune crosstalk and co-regulation between both organs. ^4,5^ Such an immune crosstalk, influenced by the interaction between host genes and the gut microbiome, might play a crucial role in the initiation of T1D.

The strongest genetic risk factor for T1D is located in the human leukocyte antigen (HLA) class II region, within the major histocompatibility complex (MHC) on chromosome 6p21. This region encodes molecules that display peptides, derived from extracellular protein antigens, to CD4^+^ T cells for their recognition. In populations with European ancestry, HLA-DQβ1 position 57 and DRβ1 positions 13 and 71 explain ∼15%, 11%, and 1% of the overall disease risk, respectively, and 80% of the risk contributed by the entire HLA region, ^6^ including HLA class I associations. ^7^ Inherited as DR-DQ haplotype pairs or diplotypes, these variants may increase or decrease predisposition to T1D up to a factor of 50 by altering the peptide-binding properties of DR and DQ (α, β) heterodimers.

HLA class II haplotypes that protect from T1D are associated with the presence of aspartic acid (D), a residue with a negatively charged side chain, at DQβ1_57_, ^8^ which is located in a key peptide-binding pocket of the DQ molecule. Conversely, haplotypes conferring near-neutral to strong susceptibility include valine (V), serine (S), and alanine (A) at DQβ1_57_, all lacking such an electrically charged side chain. The association between DQβ1_57_ and T1D is conserved in the murine ortholog of DQβ, I-Aβ. ^9^ Specific mutation of I-Aβ_57_ (S→D) in the non-obese diabetic (NOD) mouse model of T1D supports a causal role for this residue in disease by reducing the incidence of T cell-mediated islet autoimmunity. ^10^

Outside the HLA region, polymorphism of the insulin (*INS*) gene is one of the leading T1D risk factors in terms of effect size, explaining around 3% of the phenotypic variance. ^6^ Predisposing common alleles of causal variants located in the *INS* gene promoter region reduce its thymic expression, thereby increasing the frequency of anti-insulin CD4^+^ T cells in the periphery. ^11–13^ Insulin, and its precursor preproinsulin, particularly peptides from positions 9–25 of its B chain (B_9–25_), is a primary T1D autoantigen in humans and NOD mice. ^14,15^ Furthermore, the appearance of anti-insulin autoantibodies before 18 months of age is associated with the most T1D predisposing DR-DQ haplotype, DR4-DQ8. ^2,3^ Taken together, these results indicate HLA class II-restricted anti-insulin CD4^+^ T cells are responsible for the destruction of the islet β cells.

Here, we show that the overall T1D risk contributed by DRDQ diplotypes explains the thymic output of anti-insulin CD4^+^ T cell receptors (TCR). This thymic output is driven by biases during positive thymic selection that arise from charge interactions between peptides displayed by cortical epithelial cells of the thymus, T1D-associated DR and DQ β-chain residues, most notably DQβ1_57_, and TCR complementary-determining regions (CDR3) of maturing thymocytes.

However, carriers of the most T1D-predisposing diplotype, DR3-DQ2/DR4-DQ8, which encodes the highest risk DQ molecule, the transdimer (DQ2_α_, DQ8_β_), have an estimated probability of developing T1D during their life course of only 5%. Therefore, identifying environmental factors and gene-environment interactions that modulate disease risk is of critical importance not only to improve our understanding of the primary causal disease mechanisms, but also to design disease prevention interventions. During industrialization most populations have experienced a sharp increase in T1D incidence, accompanied by a decrease in the relative incidence of high-risk HLA class II diplotypes, including DR3-DQ2/DR4-DQ8, suggesting rapid changes in the prevalence of protective or predisposing environmental factors. ^16^

Different environmental factors associated with a Western lifestyle, ultimately leading to an increase in gut dysbiosis during the first months of life, could be responsible for some of these rapid epidemiological changes. Children who develop persistent islet autoantibodies and progress to T1D diagnosis typically exhibit a reduction in overall gut microbiome diversity before any autoantibody-related events, along with a decrease in the abundance of commensal bacteria from the *Bifidobacterium* genus and an increase of other phyla, ^17^ in a geographical-dependent manner. This results in a reduced capacity to metabolize human milk oligosaccharides (HMOs) and an upregulation of pathways linked to inflammation, with likely implications for the development of the host immune system. Other observational studies have identified similar patterns in children that eventually developed T1D. ^18,19^

In order to link disruptions to gut microbial ecology and the activation of anti-insulin T cells that leads to islet autoimmunity, we have modeled the similarity between insulin B_9–25_ and every protein in the gut microbiome. Our work demonstrates the existence of over 100 bacterial proteins with a statistically significant similarity to insulin B_9–25_, indicating evolutionary selection pressure to mimic insulin and a central role in the regulation of its immune tolerance.

We have identified a bacterial peptide mimic of insulin B_9–25_, displayed in the canonical B_9–23_ register, from the transketolase (TKT) enzyme family, which is highly upregulated during the transition from breastfeeding to solid food known as weaning. ^20^ This mimic is recognized by one of the few anti-insulin CD4^+^ T cells isolated from recently diagnosed T1D donor pancreata, suggesting that a targeted intervention to preserve host-microbiome symbiosis during early life could be a potential disease prevention strategy, ^4,21^ particularly in children that are genetically predisposed to T1D. ^22,23^

## Results

### HLA-DR-DQ and thymic T cell receptor selection

We analyzed the TCR α and β chain repertoires from selected donors carrying DR-DQ diplotypes in the whole protection-susceptibility risk continuum (n = 48, Table S1), with T1D odds ratio (OR) as the explanatory variable. ^24^ Purified CD4^+^ T cells (n = 349 623) from PBMCs were stimulated and loaded into a single-cell platform for preparing paired 5’ gene expression and TCR libraries. After sequencing, gene expression data analysis, and TCR assembly, we obtained 288 903 cells with both a valid gene expression profile and at least one productive TCR chain.

We partitioned the CD4^+^ T cell compartment into conventional (Tconv), recent thymic emigrant (RTE), ^25^ and regulatory (Treg) cell clusters by gene expression analysis (Figure S1 and Table S5), since these could be subject to different receptor selection constraints. As expected, the Tconv cluster was the largest (n = 258 387, clonotypes = 255 952). Owing to the low age of the cohort, this was followed by RTEs (n = 18 194, clonotypes = 18 127) and Tregs (n = 12 322, clonotypes = 12 260).

We split TCR CDR3 region sequences from α and β chains into their constituent k-mers, with k ∈ [1, 4], and aggregated counts for each (subset, chain, donor, clonotype) combination. CDR3 sequences are hypervariable and determine the peptide-binding affinity of each TCR. Counts were regressed against the log-transformed OR using a Bayesian hierarchical model to estimate the effect of DR-DQ diplotypes on each k-mer. This model takes into consideration the high variability of count data and estimates moderated effects by sharing information across variables.

We report effects as fold changes between the estimated k-mer frequencies for the most susceptible diplotype, DR3-DQ2/DR4DQ8, compared to the most protective, DR15-DQ6/DR15-DQ6. We use the false-sign rate (FSR) as an error estimate, defined as the probability that the direction of the effect, most commonly fold change, is different from the posterior median. We also report 90% credible intervals (CI) as a summary of the posterior distribution of fold changes.

For k-mers present in the CDR3β region of Tconv TCRs, those that included at least one residue with a negatively charged side chain, aspartic acid (D) and glutamic acid (E), represented most of the sequences that exhibited fold changes greater than one as T1D OR increased, at FSR ≤ 0.05 (Figure 1). Susceptible donors carrying the hydrophobic amino acid alanine (A) at DQβ1_57_ exhibited the highest frequency of D and E residues in CDR3β. Conversely, for protected donors, with at least one D at DQβ1_57_, there were fewer D and E residues in CDR3β. Instead, these residues with negatively charged side chains were replaced by amino acids of high interaction potential with D at DQβ1_57_, such as valine (V), leucine (L), and isoleucine (I).

**Figure 1:**
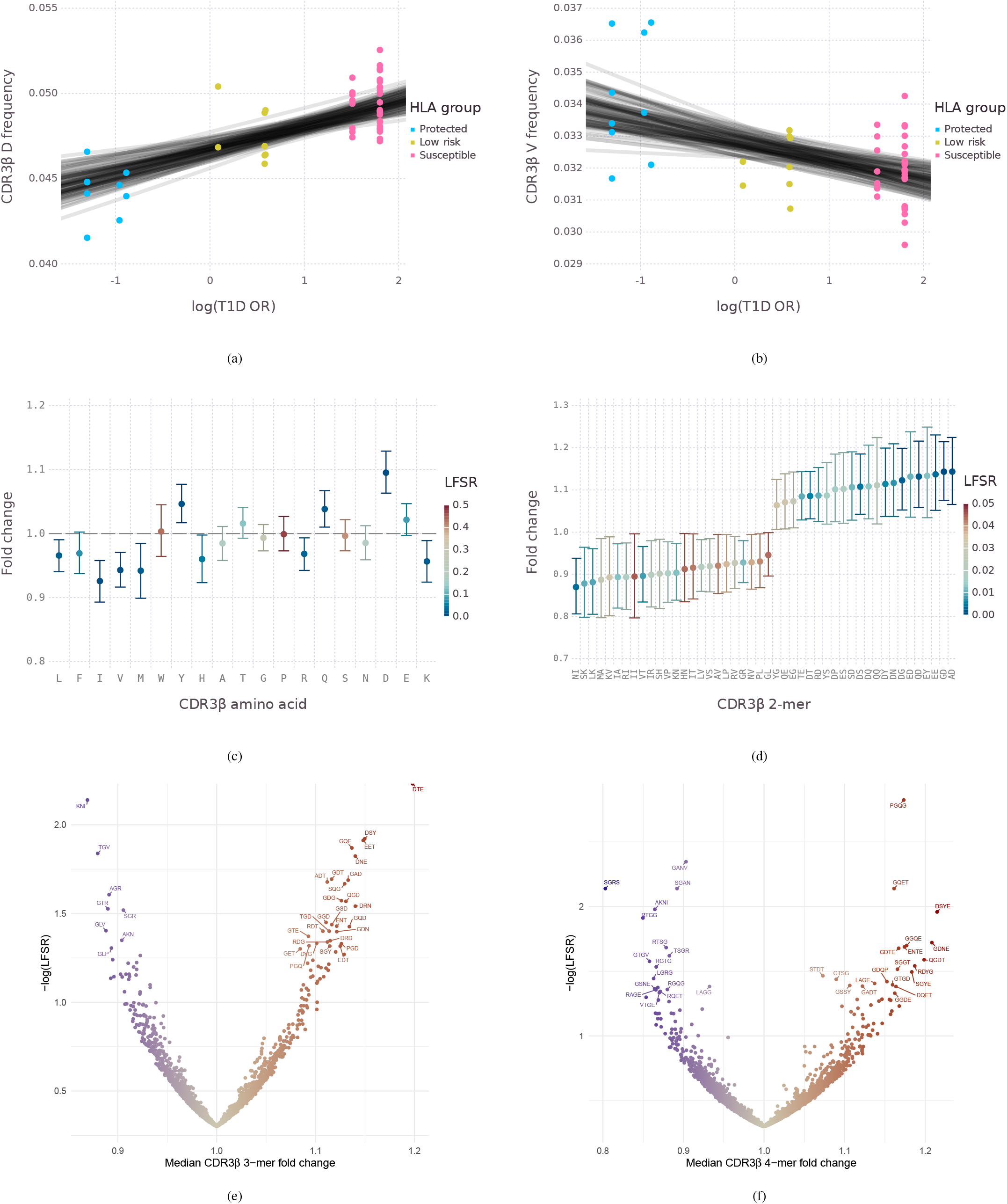
Estimates of T1D odds ratio effects, contributed by HLA DR-DQ diplotypes, on CDR3β k-mers from CD4^+^ T conventional cells. Regression lines represent 100 random draws from the posterior distribution. Error bars summarize the effects on multiple k-mers by depicting the median and 90% credible intervals of fold changes across HLA class II extremes, with the most protective DQ6 diplotypes as baseline (Table S1). Funnel plots only represent the median and the local false sign rate (LFSR). (a) Aspartic acid. (b) Valine. (c) All amino acids, or 1-mers, except cysteine. (d) 2-mers, LFSR ≤ 0.05. (e) All 3-mers with non-zero counts. 3-mers on the y axis boundary have an estimated LFSR ≈ 0. (f) All 4-mers with non-zero counts.

For RTEs and Tregs we observed the same biases in the amino acid composition of CDR3β as a function of T1D OR (Figure S2), suggesting that genetic variation effects of HLA-DR and DQ on the immune repertoire are mediated by alterations to the positive thymic selection threshold, which is common to all T cell subsets. T cell clonotypes that contain amino acids with negatively charged side chains in the CDR3β region are more likely to form a stable complex with HLA molecules expressed in the surface of thymic epithelial cells for diplotypes with a higher T1D OR as these typically lack D, a residue with a negatively charged side chain, at DQβ1_57_. For TCRα chain sequences, we also estimated systematic differences at FSR ≤ 0.05 associated with T1D risk (Figure S3). These correspond to biases in the usage of particular Vα and Jα genes, which are carried into the CDR3α region owing to the lower recombination diversity of TCRα genes. In particular, top estimated differences in k-mers match fragments of SGTYK and GGSYI, sequences encoded in TRAJ40 and TRAJ6 genes, respectively. Usage of these fragments increased in individuals carrying susceptible DR-DQ diplotypes compared to protective ones. In NOD mice, studies assessing α chain patterns have reported an expansion of anti-insulin clones with a double glycine plus serine motif (`GGS`) in the CDR3α region. ^26^.

### Negatively charged CDR3β sequences and anti-insulin responses

Next, we investigated whether these observed repertoire differences are linked to an immune response against insulin peptides. We purified activated circulating CD4^+^ T cells (n = 19 969), defined as HLA-DR^+^ CD38^+^, from children with susceptible DR-DQ diplotypes and newly diagnosed T1D (n = 5, Table S6). This activated subpopulation is enriched in autoreactive CD4^+^ T cells from patients with celiac and other autoimmune diseases. ^27^ We also sourced CD4^+^ TCR clonotype sequences (n = 1428) isolated from the islets of another group of T1D patients with susceptible DR-DQ diplotypes (n = 7, median time since diagnosis = 2 years, Table S7) from the Network for Pancreatic Organ Donors with Diabetes (nPOD). ^28^ We compared these TCR sequences against those from individuals with the highest T1D susceptibility (DR4-DQ2/DR3-DQ8, n = 23, Table S4) from our original cohort.

Both CDR3β sequences from circulating activated cells and islets had an increased frequency of D residues when compared to non-activated circulating CD4^+^ T cells from susceptible donors (Figure 2a). In the case of circulating activated cells, the posterior estimate only included fold changes above one, 90% CI = (1.115, 1.183) and FSR ≈ 0. Since few clonotypes were available from islet-infiltrating cells and our baseline was conservative, in this case the fold estimate had more uncertainty, 90% CI = (0.974, 1.120) and FSR = 0.151, but most of the posterior distribution density supported fold changes greater than one.

**Figure 2:**
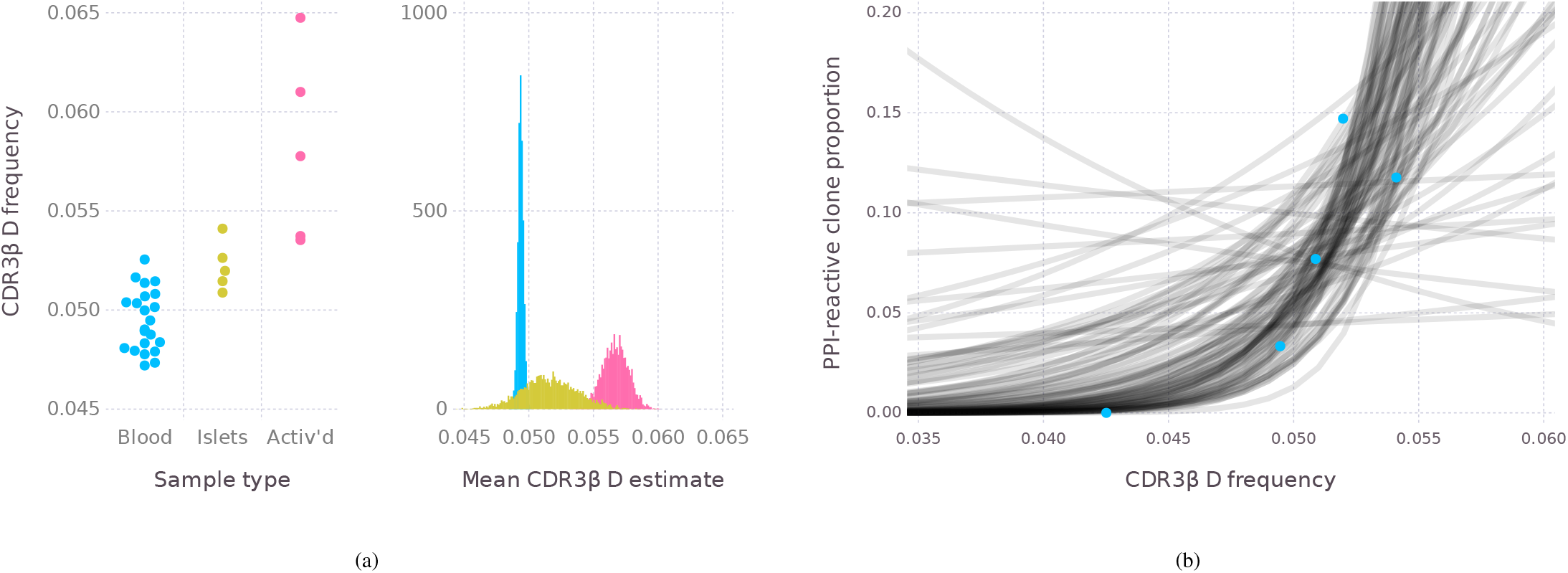
Aspartic acid (D) and anti-insulin T cell receptors. Each point denotes the summary statistics of all clonotypes in the repertoire of one donor. (a) CDR3β aspartic acid observed frequencies and posterior mean frequency estimates in CD4^+^ T cells from peripheral blood, infiltrating islets, and activated circulating cells. (B)Preproinsulin-reactive (PPI) clone proportion measured in islets as a function of CDR3β aspartic acid frequency in infiltrating CD4^+^ T cells. Regression lines represent 100 random draws from the posterior distribution.

We also analyzed TCR clonotype sequences (n = 159) from nPOD T1D donors for which reactivity against preproinsulin (PPI) peptides had been measured (n = 5, median time since diagnosis = 2 years, Table S7). ^29^ The posterior fold change in PPI-reactivity for the highest observed D CDR3β content compared to the lowest had a median = 15.36 (Figure 2b), which supports a strong effect, FSR = 0.03. The posterior credible interval covered a wide range of folds over one due to the relatively small amount of data available and the sharp increase in reactivity for high D content, 90% CI = (1.28, 320.45).

### Gut microbiome immune mimics of insulin

Differences in the proportion of anti-insulin TCRs, driven by the effects of HLA and other genetic risk variants that regulate thymic selection, may require environmental factors to cause a loss of immune homeostasis and initiate an immune response against islet β cells. Insulin mimics, encoded within microbial proteins present in the human gut, might play a fundamental role in this process. To identify these epitope mimics, or mimotopes, we investigated whether any commensal protein exhibits a statistically significant similarity to insulin B_9–25_.

We assessed the similarity between insulin B_9–25_ and any given protein as the maximum pairwise local alignment score between both sequences. This score represents the log-likelihood ratio between two models that consider amino acid substitutions as evolutionarily related or random, respectively. Log-likelihood ratios have been previously estimated using conserved regions in protein domain families, and represented as substitution matrices. ^30^ We estimated a null distribution of scores by drawing random permutations from the insulin B_9–25_ sequence and calculating pairwise local alignment scores against every reviewed entry in the human proteome.

Then, we calculated a parametric approximation to this empirical distribution and aligned insulin B_9–25_ to every protein in a gut metagenome reference catalog comprising millions of common bacterial and archaeal proteins. ^31^ This allowed the derivation of *p*-values for rare events, and false discovery rates (FDRs) to account for the large set of multiple comparisons performed. Of note, BLAST and other pairwise aligner derivatives use precalculated null distributions to reduce search runtime, and make assumptions that may not hold in case of a short query epitope. ^32^ We found over 100 microbial proteins (Figure 3) that have a significant similarity to insulin B_9–25_ at FDR ≤ 0.2, including 15 at FDR ≤ 0.05 (Table S8). The strongest association signal was found in proteins with a transketolase (TKT) N-terminal domain (Pfam PF00456). Significant associations were also present in other protein families, including YqeN DNA replication protein and hydroxyacylglutathione hydrolase (GloB). Both the number and the variety of associations suggest tolerance to insulin is regulated by a large niche of gut commensals.

**Figure 3:**
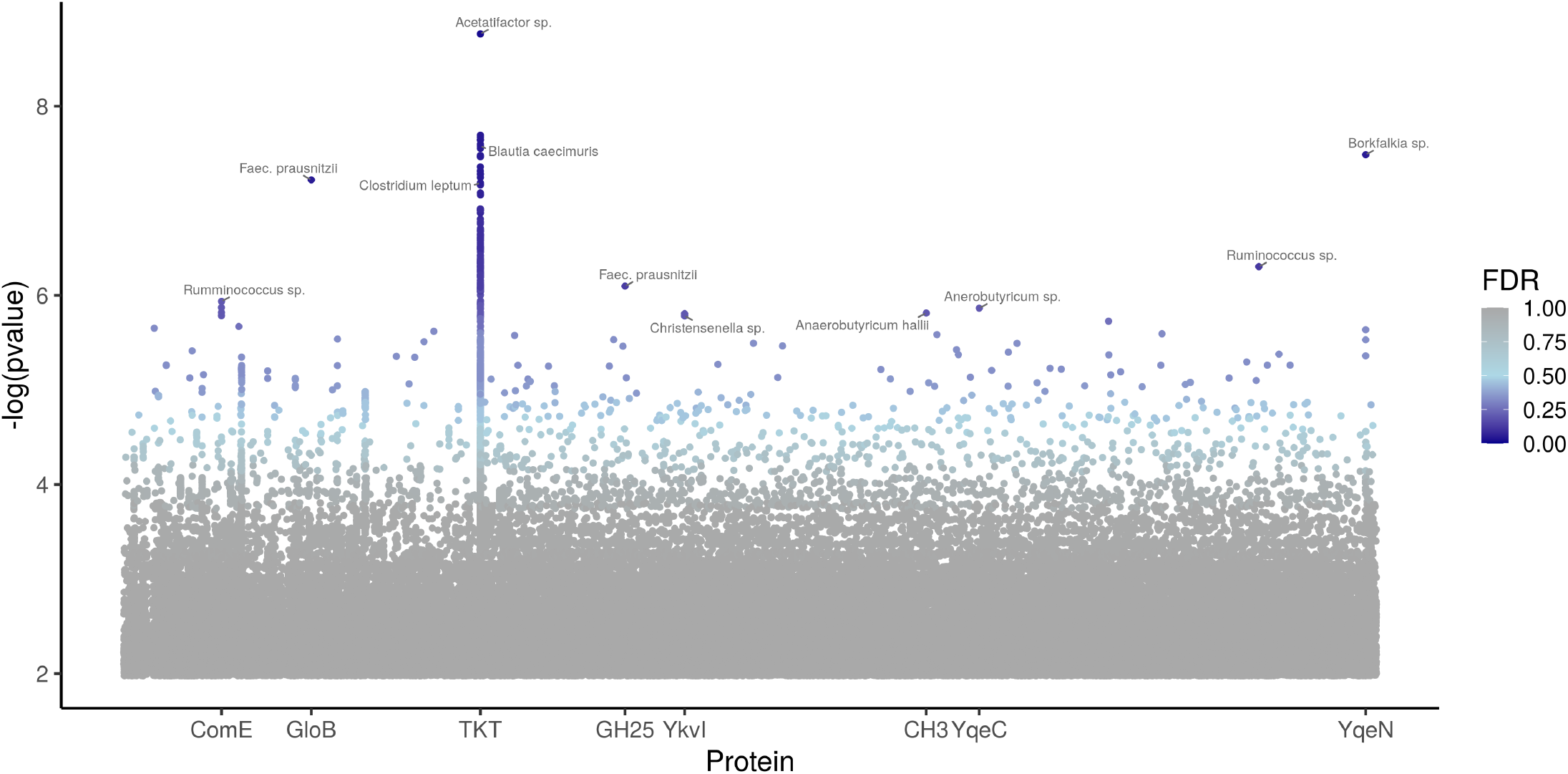
Gut microbial proteome-wide similarity with insulin B_9–25_. Each point denotes one gut microbiome protein represented against a *p*-value derived by contrasting its maximal local pairwise alignment score with insulin B_9–25_ versus a null distribution of alignment scores. Color indicates the estimated probability of false discoveries (FDR) when considering all proteins with a *p*-value lower or equal than a given threshold. Points high in the y axis denote proteins whose similarity to insulin B_9–25_ deviates from what would be expected by random chance (Table S8). Protein domains and superfamilies listed include N-terminal transketolase (TKT), yqeN DNA replication protein (YqeN), hydroxyacylglutathione hydrolase (GloB), glycosyl hydrolase family 25 (GH25), uncharacterized membrane protein (YkvI), late competence operon (ComE), selenium cofactor biosynthesis protein (YqeC), and cyclases/histidine kinases associated sensory extracellular domain 3 (CH3).

A motif analysis of the TKT N-terminal domain revealed that conserved residues, required for enzymatic function and cofactor binding, are similar or identical to those from the corresponding insulin B_9–25_ sequence (Figure S4). Therefore, this domain could have served as an ideal template for the evolution of insulin mimotopes.

### T cell cross-reactivity between microbial mimics and insulin

Functional cross-reactive bacterial mimotopes require that certain CD4^+^ T cells have TCRs that recognize both insulin B_9–25_ peptides, in the canonical B_9–23_ register, and one or more commensal epitopes. Therefore, we screened cloned CD4^+^ TCRs (n = 179), detected in islets from nPOD T1D organ donors carrying at least one copy of DQ8 (n = 6, Table S7), with TKT mimotope peptides (n = 10, Table S10) and insulin B_9–23_.

Only three islet CD4^+^ TCRs that react to insulin displayed in the B_9–23_ register are currently known (Figure 4a), including two previously reported ^33^ and one novel TCR clonotype, nPOD GSE.166H9 (Table S9). This TCR originated from a newly diagnosed patient, nPOD 6533 (DR3-DQ2/DR4-DQ8, age ∈ [0,5) years), and reacted to one of the TKT peptides (peptide 8 = GHSVEALYCILADRG) from *Clostridium leptum*. Notably, clonotype GSE.166H9 has an ET motif in the C-terminal side of its CDR3β chain, which increases in abundance proportionally to HLA class II T1D OR, FSR = 0.036.

**Figure 4:**
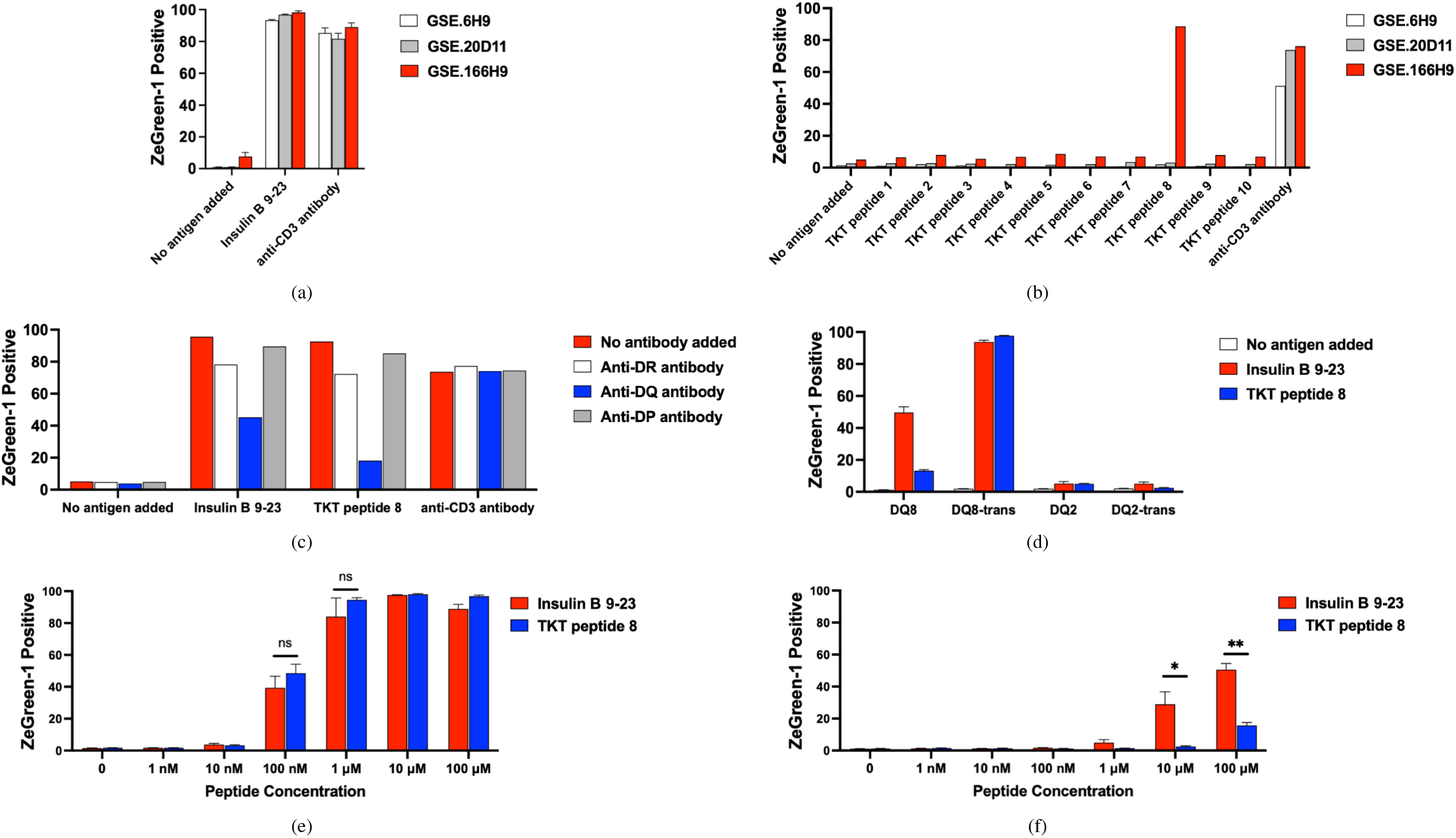
CD4^+^ T cell stimulation assays on TCRs isolated from the pancreas of T1D patients from the Network of Pancreatic Organ Donors (nPOD) and validated to recognize insulin B_9–23_. Peptide 8, which stimulates nPOD TCR GSE.166H9 (Table S9), corresponds to transketolase (TKT) from *Clostridium leptum* (Table S10). Measurements of stimulation on TCRs were obtained from 5KC T-hybridoma cells that were devoid of endogenous TCR expression and have been engineered with a ZsGreen-1 reporter gene. (a) Insulin B_9–23_ reactivity. (b) TKT epitope scan. (c) HLA class II antibodies on TCR GSE.166H9. (d) DQ presentation assay on TCR GSE.166H9. Stimulation dependent on different peptide concentrations and (e) DQ2-DQ8_trans_ or (f) DQ8 molecules. Experiments were repeated 2 (a), 3 (d), or 4 times (e and f). Mean and standard error are shown. ns: *P* > 0.05, *: *P* ≤ 0.05, and **: *P* ≤ 0.01 on a two-tailed paired t-test.

TCR GSE.166H9 recognized TKT peptide 8 and insulin B_9–23_ (Figure 4b), presented by HLA-DQ molecules (Figure 4c). This TCR responded most strongly to both peptides presented by DQ2-DQ8_trans_, consisting of DQ2α and DQ8β transdimers (Figure 4d). This TCR also recognized peptides presented by a DQ heterodimer with both chains encoded by DQ8. Intensities of responses to insulin B_9–23_ and TKT peptide 8 presented by DQ2DQ8_trans_ were similar, whereas GSE.166H9 T cells responded to insulin B_9–23_ more strongly than TKT peptide 8 when presented by DQ8.

Therefore, the most potent response was induced by both insulin B_9–23_ and peptide 8 presented by DQ2-DQ8_trans_, indicating that immune responses by TCR GSE.166H9 were initiated by these peptide-DQ complexes. The demonstration of cross-reactivity to insulin and commensal peptides, initiated by the DQ2-DQ8_trans_ molecule, helps explain why heterozygous individuals carrying both DQ2 and DQ8 have the highest predisposition to T1D.

We also sought evidence of natural processing and presentation of TKT proteins similar to those that contain putative mimotopes. We pulsed DQ6^+^ and DQ8^+^ B cell lines with a recombinant TKT protein. We then immunoprecipitated DQ and DR molecules, eluted bound peptides, and sequenced them by mass spectrometry. The top ranked eluted peptides bound to DQ6 corresponded to the insulin mimotope region (Table S11 and Figure S5). Another peptide from the mimotope region appeared to have a very similar presentation register to the only insulin B epitope that was displayed by DQ6. Bacterial TKT in the predicted mimotope area and insulin B were below detection levels in the case of DQ8. This is consistent with the properties of DQ8 peptidomes, which differ from DQ6 peptidomes in many physicochemical features ^34^ and bind insulin B_9–23_ weakly. ^35^

## Discussion

Our results support a specific biological mechanism linking two major genetic and environmental risk factors in T1D etiology: HLA class II-TCR interactions ^36^ in the thymic selection process and mimicry of insulin epitopes in the gut microbial proteome leading to increases in the frequency of T cells that can recognise both insulin and bacterial peptides, in which the interacting components have co-evolved over millions of years. Immune cross-reactivity could strengthen tolerance to an essential, highly expressed self antigen, such as insulin, as well as providing protection against the host immune system to the bacteria that encode functional insulin mimotopes and also provide essential metabolic capacity to the host.

A determinant role of positive thymic selection during interactions between DR and DQ residues, notably DQβ1_57_, and CDR3β, which alter the stability of HLA-epitope-TCR complexes, is supported by the presence of a similar bias in all three T cell subsets studied, Tconv, RTE, and Treg. These three subsets are under the same positive selection threshold in early thymocyte development. As a consequence of this bias, in case of diplotypes conferring protection, the frequency of amino acids with a negatively charged side chain in the epitope-binding CDR3β region is reduced. Conversely, in the presence of T1D-predisposing molecules such as DQ8, negatively charged CDR3β sequences are increased. These sequences have been observed to lead to highly immunogenic DQ-epitope-TCR complexes in the context of anti-gluten responses in celiac disease. ^37^

Structural models suggest CDR3 loops contribute most of the energy when TCRs form complexes with HLA and peptides, with CDR1 and CDR2 having secondary roles. ^38^ These intermolecular interactions are dominated by electrostatic forces. Contact energy models predict phenylalanine (F), isoleucine (I), and valine (V) to be favorable interaction partners of aspartic acid (D). ^39^ Conversely, interactions between aspartic acid pairs, or aspartic (D) and glutamic acid (E), are particularly unfavorable owing to charge repulsion.

Previous T1D studies employing NOD mice have observed an enrichment of amino acids with negatively charged side chains in the CDR3β region of islet T cell infiltrates. ^10,40^ These sequences typically include aspartic acid preceded by glutamine (QD). Mutation of serine at I-Aβ_57_ (S→D) provides direct support for a role of this residue in the regulation of highly immunogenic responses against insulin. ^10^ Our results from the analyses of human repertoire samples extend our understanding of why negatively charged CDR3β sequences were less frequent in the presence of D at I-Aβ_57_ in this mouse model of T1D.

Other studies have reported autoreactive TCRs are associated with the presence of hydrophobic residues in the CDR3β region. ^41,42^ This is a consequence of such residues having a high average interaction potential with other amino acids, which leads to more stable HLA-epitope-TCR complexes and influences downstream TCR signaling. However, epitopes from insulin B_9–25_ have an unusual amino acid composition, with negatively charged side chains in the Nand C-terminal regions, which suggests average trends may not be applicable to this particular case. We note that the only three available anti-insulin TCRs isolated from the pancreas of children recently diagnosed with T1D lack hydrophobic doublets at positions 6–7 of the CDR3β region, which have been described as a common pattern in self-reactive TCR repertoires.

In addition, previous studies assessing TCR α chain patterns have reported an expansion of anti-insulin clones with a double glycine plus serine motif (GGS) in the CDR3α region. ^26^ These two sequence patterns coincide with some of the k-mers that exhibit the highest fold increases as a function of T1D OR in the present study (Figures 1 and S3), consistent with HLA class II-TCR interactions increasing the frequency of anti-insulin T cells in the periphery.

Position 57 at DQβ1 is also associated with other autoimmune diseases such as celiac disease, asthma, and polyglandular autoimmunity. Position 57 in the paralog DRβ1 molecule lies in a structurally analogous pocket to DQβ1_57_, close to the epitope C-terminus and CDR3β. DRβ1_57_ has been recently fine-mapped as a strong association with several infectious and autoimmune disorders, ^43^ The existence of wide pleiotropy in these two genes, DRB1 and DQB1, is probably a consequence of the deep repertoire selection biases induced by the presence or absence of a negatively charged side chain at position 57, involving the thymic selection mechanism indicated by our results.

The existence of a higher proportion of anti-insulin TCRs in the periphery, driven solely by host genetics, is not sufficient to cause a loss of tolerance to insulin leading to a diagnosis of T1D. This is evidenced, for example, by the continuous increase of T1D incidence in multiple geographical locations that cannot be explained by changes in the gene pool, and suggests alterations to some environmental factors. ^16^

Molecular mimicry of host antigens has been widely explored as a causal mechanism of common autoimmune diseases mediated by environmental factors. In the case of T1D, early efforts were mainly centered around enteroviral mimics of a primary islet autoantigen, glutamate decarboxylase 65 (GAD65). ^44,45^ Possible instances of insulin B chain mimicry by microbial antigens, with varying levels of support, have been reported. These include environmental bacteria that cross-react with human tetramer-sorted CD4^+^ T cells, ^46^ a gut bacterial antigen that cross-reacts with a NOD CD4^+^ clonotype, ^47,48^ and fungal sequences that cross-react with a human CD8^+^ clonotype. ^49^ Other reported findings involve the insulin signal peptide ^50^ and defective ribosomal products (DRiPs) as a target antigen. ^51^

The degeneracy of TCRs, meaning some clonotypes can crossreact with thousands or even millions of different peptides, ^52^ has led to questions about the validity of some evidence and the overall role of mimicry in disease pathogenesis. For example, islet-specific CD8^+^ T cells have been reported to gain effector function from by-stander activation, and not molecular mimicry. ^53^ Generating high-grade evidence is further complicated by the paucity of islet-specific human clones available. ^54^

By initially framing this question as a statistical sequence analysis problem, we were able to circumvent some of these issues, and reject the null hypothesis of a lack of evolutionary relatedness between more than 100 bacterial sequences and insulin B_9–25_. This suggests symbiotic co-evolution between hosts and commensals, ^55^ and a role in the regulation of immune tolerance to insulin. We have previously reported analogous results using a Bayesian mixture model. ^22^

We obtained functional evidence for this mimicry by demonstrating cross-reactivity to one of the only three identified antiinsulin DQ-restricted TCRs isolated so far from the pancreata of recently diagnosed T1D patients. Our findings draw parallels to other emerging autoimmune disease mechanisms driven by epitope mimicry, such as those observed in systemic lupus erythematosus, multiple sclerosis, rheumatoid arthritis, and narcolepsy. ^56^

Some of the identified microbial commensals, which harbor sequences more similar to insulin B_9–25_ than what would be expected by random chance, have been previously associated with T1D in a variety of longitudinal and cross-sectional studies. *Clostridium leptum*, which encoded the TKT mimotope that stimulated the TCR from an islet-resident T cell (Figure 4), was diminished in NOD mice that progressed to T1D. ^57^

A fecal transplant trial in adults with active T1D halted the disease in some patients. *C. leptum* abundance increases were among the predictors of response to the transplant. ^58^ *C. leptum* also exhibited the strongest correlation with certain plant-based food diets, consisting of maltose, sucrose, starch, and other carbohydrate sources, in a large observational study that profiled gut microbiomes. ^59^

TKT mimics contain a N-terminal transketolase domain and exhibit sequence similarity to a few members of the apulose-4phosphate transketolase enzyme subfamily. These are involved in the catabolism of D-apiose, found in pectins, which could point to imbalanced plant-based nutrients in the development of gut dysbiosis during early life. Diets with pectins increased the ratio of Bacteroidota (Bacteroidetes) to Bacillota (Firmicutes) in the gut and promoted T1D in NOD mice. ^60^

Bacterial TKT is among the most highly upregulated commensal enzymes during the first year of life, ^20^ probably to aid processing of incoming fiber and other carbohydrates from solid foods. Successive colonization waves, particularly at the time of weaning, which precedes the peak of insulin autoantibody positivity between 9 and 18 months of age, represent a period of physiological stress when immune tolerance to a myriad of antigens is facilitated by interactions with the microbiome.

Further studies are required to determine the role of insulin mimics in the maintenance of immune homeostasis and susceptibility to disease. Gut microbiome dysbiosis, increased gut permeability, and chronic low-grade inflammation, owing to a myriad of modern lifestyle factors, ^61–63^ may interfere with the role of commensal mimics in the induction of sufficient insulin-reactive Tregs. ^64^

These environmental factors could also lead to an excessive growth of certain commensal species that encode insulin mimics causing the expansion of insulin-reactive Tconvs, the number of which is increased in carriers of susceptible HLA class II diplotypes, as shown here. These results and other associations between the microbiome and T1D have motivated a phase II clinical trial that is investigating whether a daily probiotic supplementation may reduce the incidence of islet autoimmunity. ^23^

## Data Availability

All data produced in the present study will be available upon reasonable request to the authors.

## Acknowledgements

We thank all volunteers participating in this study. We are grateful to the study volunteers and staff associated with JDRF Diabetes, Genes, Autoimmunity and Prevention (D-GAP), including the Wellcome Clinical Research Facility (Addenbrooke’s Clinical Research Centre, Cambridge, UK) and Investigating Underlying Causal Mechanisms in Type 1 Diabetes (DMech) hospital sites, including the Institute of Mother and Child (Warsaw, Poland). We thank Network for Pancreatic Organ Donors with Diabetes (nPOD) and their families.

We thank Aaron Michels (Barbara Davis Center for Childhood Diabetes, University of Colorado School of Medicine, Aurora, Colorado, USA) for helpful discussions and for providing EBV cell lines expressing various HLA class II molecules, and Wojciech Szypowski (Polish Society for Autoimmune Diseases, Warsaw, Poland) for sample logistics. We thank Nicola Burgess-Brown and Alejandra Fernández Cid (University of Oxford, Oxford, UK) for helpful discussions and production of recombinant microbial proteins. We thank members of the JDRF/Wellcome Diabetes and Inflammation Laboratory (DIL, University of Oxford): Heather McMurray, Shannah Donhou, Sarune Kacinskaite, Michael Ellis, Sandra Banks, Georgina Burton and past members at the University of Cambridge (Cambridge, UK) led by Helen Stevens for blood sample processing. We thank Raqeeb Mahmood and Sylwia Kopijasz for managing ethics and blood donor recruitment; Hong Harper for managing the patients’ registry and DGAP data; Florent Yvon for HLA imputation of SNP data from D-GAP patient DNA; Jamie Inshaw for information concerning D-GAP donor metadata; Olga Platonova for managing finance and funding; Moustafa Attar (Oxford Genomics Centre, University of Oxford) for assistance with single cell sequencing; and Holly Roach for contributions to the manuscript.

This research was performed with the support of nPOD (RRID:SCR_014641), a collaborative T1D research project. The content and views expressed are the responsibility of the authors and do not necessarily reflect the official view of nPOD. Organ procurement organizations (OPO) partnering with nPOD to provide research resources are listed at https://www.jdrfnpod.org/for-partners/npod-partners. This work has been supported by a JDRF (4-SRA-2017473-A-N) and Wellcome (107212/A/15/Z) Strategic Award to JAT and LSW. D-GAP was a centre grant funded by the JDRF (1-2007-1803) to Mark Peakman, Tim Tree, JAT, LSW, Polly J. Bingley, and David B. Dunger. ARG was also supported by the DARPA Probabilistic Programming for Advanced Machine Learning (PPAML) program. The work performed in the University of Colorado has been supported by the National Institutes of Diabetes and Digestive and Kidney Diseases (R01DK099317, R01DK032083, P30DK116073), and JDRF (5-SRA-2018-557-Q-R to nPOD).

We dedicate this work to the memory of Dr Robert Goldstein MD (1941– 2018), Professor David B. Dunger FRCP FMedSci (1948–2021), and Professor Hugh O. McDevitt MD ForMemRS (1930–2022).

## Author contributions

ARG, JAT, and MLP designed and co-led the study. ARG conceived and developed TCR repertoire and epitope mimicry statistical models. ARG, MLP, and JAT drafted the manuscript. MN contributed data, comments on manuscript drafts, and designed and produced the T cell cross-reactivity results with the assistance of LGL and AMA. MLP and AP led experiments with help of ML, HS, and LGL. AP, DT, MLP, ARG, JAT, and LSW selected HLA class II diplotypes. RF, MLP, AP, and ML performed additional single-cell BD Rhapsody experiments. AP and LSW supervised QC of D-GAP’s DNAs for iChip and HLA genotyping. DT processed BD Rhapsody single-cell data and genotype data queries. RPD and NT conducted immunopeptidomics experiments and mass spectrometry data analysis. LSW, JAT, MLP, and AS designed the clinical cohort collections. LT contributed to the planning of the study and discussed the results. CLS was responsible for ethics and sponsor approvals. All authors approved the final manuscript.

## Data and code availability

All raw data and source code will be deposited in public repositories ahead of publication following FAIR guidelines. Pre-processed data will also be made available to ease reproducibility.

## Methods

### Method details

#### Circulating CD4^+^ T cell repertoires

##### Donor selection

Donors were sourced from the JDRF Diabetes, Genes, Autoimmunity, and Prevention (D-GAP) cohort. Donors were selected to cover the whole protection-susceptibility risk continuum (Table S1), according to T1D risk contributed by HLA DR-DQ diplotypes. ^24^ London Hampstead Research Committee of the NHS Health Research Authority gave ethical approval for this work, ref. no. 08/H0720/25. Upon closure of the D-GAP study, samples were transferred to Investigating Genes and Phenotypes of Type 1 Diabetes, Cambridgeshire 2 Research Ethics Committee, ref. no. 08/H0308/153.

##### Genotyping

Genomic DNA was prepared from PBMCs or whole blood using QiaAmp DNA Blood kit (Qiagen), or phenolchloroform extraction. Haplotypes were measured using Taqman sequencing (Applied Biosystems) of four SNPs (rs2187668, rs660895, rs9271366, and rs7454108), complemented with RELI SSO (DYNAL Biotech) classical HLA typing. HLA class II types were further confirmed with ImmunoArray-24 BeadChip v2.0 (Infinium) or HumanImmuno BeadChip v1.0 (Illumina), and HLA imputation, ^65^ along with further SSP classical HLA class II typing (MC Diagnostics and Oxford Transplant Centre).

##### PBMC processing

PBMC isolation, cryopreservation, and thawing were performed as previously described. ^25^ PBMCs were isolated using Lympholyte (CEDARLANE). PBMCs were cryopreserved in heat-inactivated filtered human AB serum (Sigma-Aldrich) and 10% DMSO Hybri-MAX (Sigma-Aldrich) at a concentration of 2–10 × 10^6^ / ml, and were stored in liquid nitrogen. PBMCs were thawed in a 37 °C water bath for 2 min, and then washed by adding 1 ml of AB serum to cells, followed by adding 10 ml of cold (4 °C) X-VIVO (Lonza), containing 10% AB serum per up to 1 × 10^7^ cells, both in a drop-wise fashion. PBMCs were then washed again with 10 ml of cold (4 °C) X-VIVO containing 1% AB serum per 1 × 10^7^ cells.

##### CD4^+^ cell purification

CD4^+^ T cells were purified from thawed PBMCs using negative selection with EasySep Human CD4^+^ T Cell Enrichment Cocktail (STEMCELL Technologies), then washed with X-VIVO medium (Lonza) containing 5% human AB serum, and resuspended at 1 × 10^5^ cells / well (96 well plate) in a final volume of 200 µl X-VIVO medium (Lonza) containing 5% human AB serum. Cells were activated with PMA/ionomycin (eBioscience) for 2 h, then harvested, washed, resuspended in PBS, and counted.

##### Single-cell sequencing

CD4^+^ T cells were washed in PBS with 0.04% BSA and re-suspended them at a concentration of 800–1200 cells / µl, before capturing ∼5 × 10^3^ single cells in droplets using the Chromium platform (10x Genomics). Generation of paired gene expression and TCR libraries was performed using the Chromium Single Cell V(D)J Reagent Kits v1 and v1.1b. Quantification of libraries was carried out using Qubit dsDNA HS Assay Kit (Life Technologies) and D1000 ScreenTape (Agilent). Libraries were sequenced on HiSeq 4000 and NovaSeq 6000 (Illumina) to achieve an average of 2 × 10^4^ read pairs / cell for gene expression libraries, and 5 × 10^3^ read pairs / cell for TCR libraries.

##### Single-cell data preprocessing

Single-cell RNA and TCR sequencing libraries were processed separately using Cell Ranger v4.0.0 (10x Genomics) to obtain gene counts and receptor assemblies. These were then merged into a single gene expression matrix and a single TCR database, which mapped gene counts and TCR chains to unique molecular identifiers and donor barcodes.

#### Circulating activated CD4^+^ T cell repertoires

##### Donor selection

Donors were sourced from the study Investigating Underlying Causal Mechanisms in Type 1 Diabetes (DMech). Donors were selected among those that had been recently diagnosed and carried a susceptible HLA DR-DQ diplotype (Table S6), conferring a T1D OR > 0.9 in log_10_ scale. Donors were genotyped as described in the previous section. South Central Oxford A Research Ethics committee of the NHS Health Research Authority gave ethical approval for this work, ref. no. 18/SC/0559.

##### CD4^+^ cell purification

Fresh whole-blood samples from newly diagnosed T1D patients were shipped overnight at room temperature. From each donor, CD4^+^ T cells were isolated from 10 ml of blood using RosetteSep (STEMCELL Technologies) according to the manufacturers’ instructions. Negatively selected CD4^+^ T cells were washed with PBS plus 2% FBS.

##### Fluorescence-activated cell sorting

Cells were incubated with the following fluorochrome conjugated antibodies: CD38-BV421 (Biolegend), HLA-DR-AF700 (Biolegend), CD3-BV510 (BD Biosciences), and CD4-BUV395 (BD Biosciences) in Brilliant Stain Buffer (BD Biosciences). Following incubation for 30 min at 4 °C, cells were washed two times and resuspended in PBS + 1% FBS for cell sorting, at 4 °C in a BD FACSAria Fusion sorter (BD Biosciences). CD3^+^ CD4^+^ HLA^-^ DR^+^ CD38^+^, control pools of CD3^+^ CD4^+^ HLA-DR^-^, and CD3^+^ CD4^+^ HLA-DR^+^ CD38^-^ were FACS-purified (2–3.5 × 10^4^ cells / donor), washed, and processed for BD Rhapsody Express Single-Cell analysis (BD Biosciences). FACS-sorted cells were incubated with different oligo-conjugated sample barcoding antibodies (Sample Multiplexing Kit, BD Biosciences) for 20 min on ice.

##### Cell incubation

Barcoded cells from each batch of donors (total three batches) were then pooled together in a 5 ml FACS tube (Falcon) and washed in cold PBS + 2% FBS. Cell pools were then incubated for 5 min at 4 °C with Human Fc block (BD Biosciences) and then immediately incubated with a mastermix of 62 oligo-conjugated AbSeq antibodies (BD Biosciences) ^66^ for 45 min on ice. Following AbSeq incubation, cells were washed three times in cold BD Sample Buffer (BD Biosciences), to remove any residual unbound antibody, filtered and resuspended in 620 µl of cold BD Sample Buffer for cell capture. Each of the three patient pools was loaded on a BD Rhapsody cartridge (BD Biosciences), and we aimed to retrieve approximately 2 × 10^4^ cells / pool.

##### Library preparation

mRNA, TCR, AbSeq, and sample tag cDNA products were initially amplified for 11 cycles (PCR1). The resulting PCR1 products were purified by double-sized selection using AMPure XP magnetic beads (Beckman Coulter), to separate the shorter AbSeq (∼170 bp) and sample tag (∼250 bp) products from the longer mRNA (350–800 bp) and TCR (600–1000 bp) products. The purified mRNA (10 cycles), sample tag (10 cycles) and TCR (15 cycles) PCR1 products were then further amplified using their respective nested PCR primer panels (PCR2) on separate reactions. The resulting mRNA, sample tag, and TCR PCR2 products were purified by size selection. The concentration, size, and integrity of the PCR products was assessed using both Qubit High Sensitivity dsDNA kit (ThermoFisher Scientific) and the Agilent 4200 Tapestation system High Sensitivity D1000 (Agilent). The final products were normalized to 2.5 ng / µl (mRNA), 1 ng / µl (sample tag and AbSeq), and 0.5 ng / µl (TCR). These underwent a final round of amplification (6 cycles for mRNA, sample tag, and AbSeq; 7 cycles for the TCR libraries) using indexes for Illumina sequencing to prepare the final libraries. Final libraries were quantified using Qubit and Agilent Tapestation and pooled (25% mRNA : 14% TCR : 57% AbSeq : 4% sample tag ratio) to achieve a concentration of 5 nM.

##### Single-cell sequencing

Single-cell capture and cDNA library preparation, including TCR libraries, was performed using the BD Rhapsody Express Single-Cell analysis system (BD Biosciences), using the VDJ CDR3 protocol to generate the mRNA, TCR, AbSeq, and sample tag libraries. The targeted mRNA panel used in this assay was based on the pre-designed Human T-cell Expression primer panel (BD Biosciences), combined with a custom designed primer panel (containing an additional 306 primer pairs), as previously described. ^66^ Final pooled libraries were spiked with 15% PhiX control DNA to increase sequence complexity and were sequenced (75 bp, 225 bp paired-end) on a NovaSeq system (Illumina).

##### Single-cell data preprocessing

Single-cell RNA and TCR sequencing libraries were processed jointly using the cloud-based BD Rhapsody Targeted Analysis Pipeline (Seven Bridges Genomics). A notable difference with other platforms is the absence of a N-terminal cysteine in CDR3 chain calls, which needs to be considered when counting k-mers in downstream data analysis.

#### Infiltrating CD4^+^ T cell repertoires

##### Donor selection

Donors were sourced from the Network for Pancreatic Organ Donors with Diabetes (nPOD, Table S7). Studies involving human participants were reviewed and approved by the University of Florida Institutional Research Board, ref. no. IRB201600029.

##### Peptide elution from HLA-DQ molecules

##### Protein production

Recombinant transketolase (TKT) protein was made by the Centre for Medicines Discovery (University of Oxford). Recombinant TKT from *B. caecimuris*, Integrated Gene Catalog (IGC) ^67^ entry MH0370_GL0036213, was produced in *Escherichia coli* BL21(DE3)-pRARE2, then purified by nickel affinity chromatography and size exclusion chromatography with endotoxin removal. Among all our hits in MGnify, ^31^ *B. caecimuris* was the first entry in IGC, a large cohort assembly with estimated abundances, and present in more than 10% of the individuals. IGC entry MH0370_GL0036213 corresponds to MGnify entry MGYG000164756_00723, with the latter lacking a nine amino acid sequence in the N-terminus, probably arising due to an alternative transcription start site prediction.

##### Cell culture and pulsing

Peptide elution from HLA-DQ molecules was performed as previously described. ^68^ EBV cells were grown to a total cell count of 1 × 10^8^ in complete RPMI (R10). Spent R10 was removed, and cells were incubated with 500 µg of TKT or insulin (Biomm SA) in 5 ml of R10 for 2 h at 37 °C. R10 was added to full plate volume (15 ml) and cells further incubated at 37 °C for 18 h. Cells were collected, pelleted (400 × g, 5 min) and washed with PBS. Washed cells were pelleted again and frozen at −20 °C for further use.

##### Immunoprecipitation

Harvested pellets were washed in PBS then lysed by mixing for 30 min with 2 ml of lysis buffer (0.5% v/v IGEPAL 630, 50 mM Tris pH 8.0, 150 mM NaCl) and 1 tablet Complete Protease Inhibitor Cocktail EDTA-free (Roche) / 10 ml buffer at room temperature. Lysate was clarified by centrifugation at 1000 × g for 10 min followed by a 20 000 × g spin step for 45 min at 4 °C. 2 mg of anti-HLA-DQ SPVL3 antibody-PAS was incubated with lysate overnight with gentle rotation at 4 °C. Resin was collected by gravity flow, and flow-through lysate was collected for sequential incubation with IVA12-PAS. Antibody–resin–HLA complexes were sequentially washed (15 ml 0.005% IGEPAL, 50 mM Tris pH 8.0, 150 mM NaCl, 5 mM EDTA, 15 ml of 50 mM Tris pH 8.0, 150 mM NaCl, 15 ml of 50 mM Tris pH 8.0, 450 mM NaCl, and 15 ml of 50 mM Tris pH 8.0), and 5 ml of 10% acetic acid was used to elute bound HLA-DQ complexes from the PAS-antibody resin.

##### High-performance liquid chromatography

Elutions were vacuum centrifuged for drying, dissolved in loading buffer (0.1% v/v trifluoroacetic acid [TFA], 1% v/v acetonitrile in water), injected by an Ultimate 3000 HPLC system (ThermoFisher Scientific), and separated across a 4.6 mm × 50 mm ProSwift RP-1S column (ThermoFisher Scientific). Peptides were eluted using a 1 ml / min gradient over 5 min from 1–35% acetonitrile in 0.1% TFA and fractions were collected every 30 s for 18 fractions. Peptide fractions 1–12 were combined into odd and even fractions, then dried.

##### Mass spectrometry

HPLC fractions were dissolved in loading buffer and analyzed by an Ultimate 3000 HPLC system coupled to a high field Q-Exactive (HFX) Orbitrap mass spectrometer (ThermoFisher Scientific). Peptides were initially trapped in loading buffer, before RP separation with a 60 min linear acetonitrile in water gradient of 2–35% across a 75 µm × 50 cm PepMap RSLC C18 EasySpray column (ThermoFisher Scientific) at a flow rate of 250 nl / min. An EasySpray source was used to ionise peptides at 2 × 10^3^ V, and peptide ions were introduced to the MS at an on-transfer tube temperature of 305 °C. Ions were analyzed by data-dependent acquisition. Initially a full-MS1 scan (1.2 × 10^5^ resolution, 60 ms accumulation time, AGC 3 × 10^6^) was followed by 20 data-dependent MS2 scans (6 × 10^4^ resolution, 120 ms accumulation time, AGC 5 × 10^5^), with an isolation width of 1.6 m / z and normalized HCD energy of 25%. Charge states of 2–4 were selected for fragmentation.

#### CD4^+^ T cell stimulation assay

##### T-hybridoma cells

TCR sequences were identified from CD4^+^ T cells in the islets or pancreas slices of T1D organ donors carrying the DR4-DQ8 haplotype distributed by nPOD: nPOD 69, 6323, 6342, 6367, 6472, and 6533 (Table S7), as described previously. ^69^ A total of 171 TCRs were expressed in 5KC T-hybridoma cells, devoid of endogenous TCR expression and engineered with a ZsGreen-1 reporter gene preceded by the nuclear factor activated T cells (NFAT) binding sequences. ^70^ TCR-expressing 5KC cells (2 × 10^4^ cells / well) were cultured with 100 µM or designated concentrations of peptides in the presence of antigen presenting cells, Epstein-Barr virus-transformed autologous B cells (1 × 10^5^ cells / well) or K562 cells transduced with each designated HLA class II molecule ^69^ (5 × 10^4^ cells / well) in round-bottom 96-well plate for 16–22 h, as described in figure legends.

##### Peptide stimulation

Peptides at > 95% purity were purchased from Genemed Synthesis (Table S10). Cultures with and without anti-CD3*ε* antibody at 5 µg / ml (clone 125-2C11) were included in each assay as positive and negative controls, respectively. To determine which HLA molecules were presenting antigen to T cells, anti-HLA-DR (clone L243/G46-6), anti-HLA-DQ (REA303), or anti-DP (B7/21) antibodies were added at 12.5 µg / ml.

#### Quantification and statistical analysis Single-cell data analysis

##### Quality control

We called cells from unique molecular identifiers with a minimum of 300 genes expressed. We also removed genes not present in at least 50 cells to keep the expression matrix tractable. Furthermore, we applied batch-dependent cutoffs to remove outliers suspected to be cell doublets or multiplets. We also filtered cells with more than 15% of mitochondrial expression to discard those undergoing apoptosis. We only retained clonotypes originating from a consensus assembly and productive rearrangements.

##### Normalization

After data cleanup, we normalized all expression values to 10^4^ reads / cell and applied a log_10_ transformation. Next, we discarded all but the top 5 × 10^3^ most variable genes, and regressed out differences due to sequencing depth and mitochondrial expression using a generalized linear model.

##### Community detection

We aligned cells from all samples using batch-balanced nearest neighbors, ^71^ reduced the dimensionality to 2D, ^72^ called clusters, ^73^ and performed multivariate differential expression to detect subpopulation markers (Table S5). ^74^

##### k-mer quantification

We excluded a N-terminal prefix of 4 residues and a C-terminal suffix of 3 residues from each CDR3 to avoid the HLA class II binding bias that constrains gene usage. Subsequently, we computed k-mer sequence frequencies by aggregating clonotype counts for each combination of k-mer length, CD4^+^ subpopulation, and donor.

#### Bayesian inference on CD4^+^ T cell repertoires

##### Markov chain Monte Carlo inference

We drew samples from posterior distributions using the No-U-Turn sampler (NUTS) implemented by Stan v2.32.1. ^75,76^ NUTS was run with default control parameters. In case of the model defined to estimate the effects of HLA on TCR repertoires (Generative model 1), for efficiency reasons, we removed covariates age, sex, autoantibody status, and T1D status since these had little predictive value. We also re-ran inference excluding autoantibody-positive and T1D donors, obtaining analogous differential expression results.

##### Model checking and diagnostics

We used the split-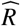 convergence diagnostic statistic to flag chain autocorrelation ^77^ issues. For all inference runs, 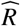 deviated from 1 by less than 0.05, the standard convergence bound. We also carried posterior predictive checks with Bayesian *p*-values on 50 CDR3β 2-mers with the lowest and highest LFSR using sample mean as a test statistic. ^78^ No tail-area probabilities were near 0 or 1, which indicated replicated datasets were similar to observed data and therefore no major model failures could be identified.

**Generative model 1:**
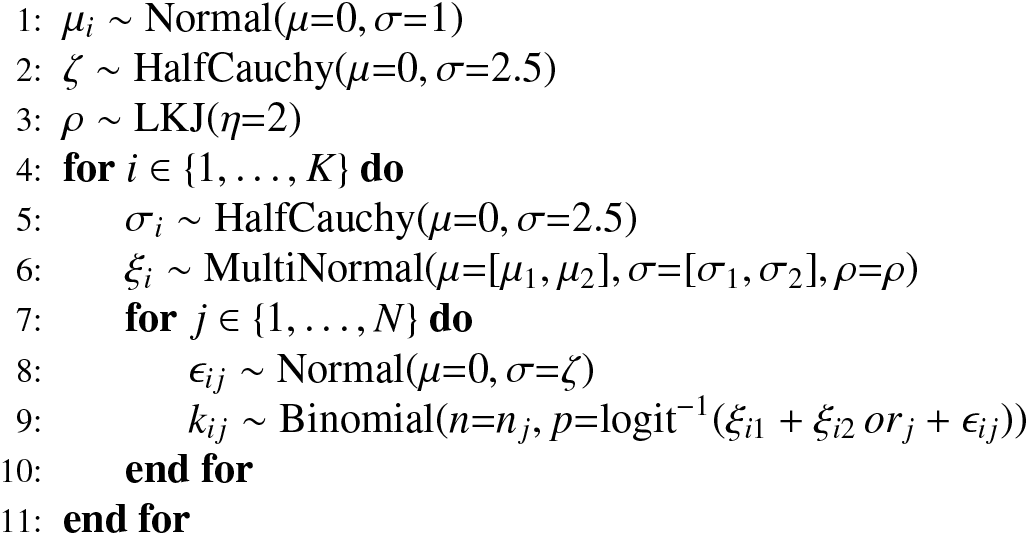
Hierarchical mixed HLA effects model. Observed counts *k*_*i j*_ on k-mer *i* and donor *j*, out of a total *n* _*j*_ k-mers, are used to estimate the effect *ξ*_*i*_ explained by the T1D log-odds ratio *or*_*j*_ in a regression with normally distributed random effects. Regression intercepts, slopes, intercept-slope covariances, and random effects are shrunk by sharing information across k-mers with a hierarchical model.

**Generative model 2:**
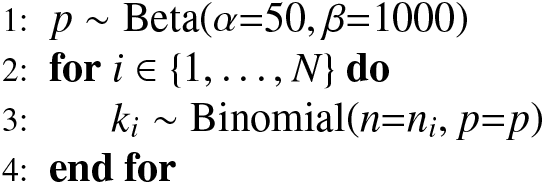
Beta-binomial aspartic acid proportion model. Observed aspartic acid counts *k*_*i*_ on donor *i*, out of a total *n*_*i*_ amino acid counts, are used to estimate the true hidden proportion *p* on each set of group observations. The prior belief for the frequency of aspartic acid *p* is modeled with an informative prior, using the posterior inferred previously (Generative model 1).

**Generative model 3:**
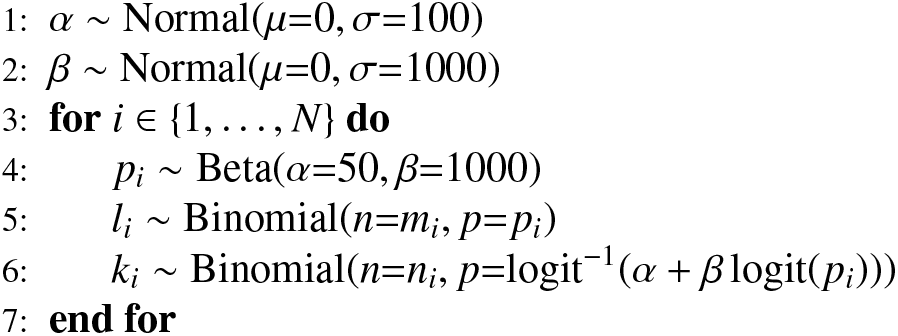
Binomial preproinsulin-reactivity (PPI) regression model. Observed aspartic acid counts *l*_*i*_ on donor *i*, out of a total *m*_*i*_ amino acid counts, are used to regress the number of PPI clonotypes *k*_*i*_, out of a total *n*_*i*_ clonotypes tested, using a measurement error structure. The prior belief for the frequency of aspartic acid *p* is modeled with an informative prior, using the posterior inferred previously (Generative model 1).

#### Insulin mimotope discovery

##### Alignment score model

We can formulate the problem of finding probable mimotopes to an epitope of interest as a statistical inference problem where we have to identify pairwise similarity that exceeds what would be expected by random chance. ^79^ We measured similarity as the maximum local pairwise alignment score between two protein sequences, computed using the Smith-Waterman algorithm. ^80^ We used a cost model defined by infinite gap penalties and a BLOSUM 80 substitution matrix. ^30^

##### Empirical null distribution

We drew 10^5^ random permutations of the epitope of interest, insulin B_9–25_. On each iteration, we performed a pairwise local alignment against all 20 375 reviewed canonical protein isoforms from *Homo sapiens* in the UniProt database. This yielded more than 2 × 10^9^ scores.

##### Non-parametric inference

Equipped with an empirical null distribution, it is straightforward to assess the significance of maximal local pairwise alignment scores of the non-permuted epitope against any set of proteins, e.g. all predicted open-reading frames from a gut microbiome assembly. This is simply done by calculating one-sided *p*-values, or evaluating the complementary cumulative distribution function (ccdf) for each score.

##### Parametric inference

Since typically only the highest alignment score from each pairwise alignment is considered, we want to model the statistical behavior of the maximum of a sequence of independent and identically distributed random variables, all scores for a given pairwise alignment, *max*{*S* _1_, …, *S* _*n*_}. The Fisher-Tippett-Gnedenko or extremal types theorem states such a maximum, after proper renormalization, can only converge to the generalized extreme value (GEV) distribution. The cumulative distribution function, defined on the set {*z* | 1 + *ξ*(*z* − *µ*)*/σ >* 0}, is given by

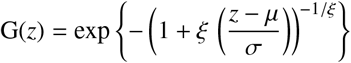

##### Maximum likelihood estimation

We can fit a GEV distribution to the observed data by maximum likelihood. This maximum likelihood fit can be obtained by solving the constrained optimization problem defined below, and is guaranteed to exist and be unique. ^81^ We used an interior point line search filter algorithm ^82^ to estimate such a fit to a set of maximal alignment scores and calculate a parametric approximation to the null distribution of local alignment scores. The GEV log-likelihood to maximize is

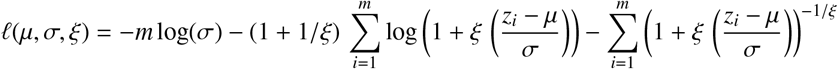

subject to the constraint

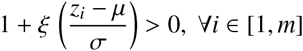

If *ξ* = 0, we require separate treatment

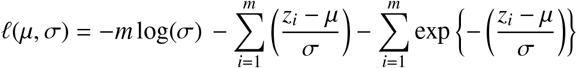

We have previously identified many of the same insulin mimotopes using a different generative model-based method. ^22^ Other approaches to the same problem have relied on human-curated results from BLAST searches performed on fully assembled genomes from cultured species. ^46,48^ Consequently, selected peptides lack statistical significance or come from organisms that do not have humans as a host.

##### Sequence motif conservation

*Amino acid information content*. The information content is defined as the reduction in entropy after some message is received. In case of a protein motif, the information content *I* at position *i* is given by the difference in entropy between a uniform distribution of 20 amino acids and the observed distribution

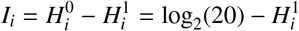

where the entropy *H* of residue *i* is given by

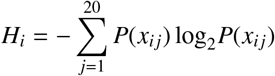

##### Consensus motif logo

We built a consensus motif logo for N-terminal transketolase (TKT) in the region of insulin mimicry by representing the amino acid distribution at each position *i*, rescaled by its information content *I*_*i*_ (Figure S4). ^83^ Since the profile hidden Markov model (pHMM) for TKT (Pfam PF00456) in the region of interest has near zero probability of insertions or deletions, this allowed establishing a one-to-one mapping between pHMM states and residues.

#### Liquid chromatography and mass spectrometry

##### Peptide identification

Raw data files were analyzed with PEAKS X (Bioinformatic Solutions) using a protein sequence database containing 20 606 reviewed human entries from UniProt, supplemented with the sequence of *B. caecimuris* transketolase protein (Figure S5). No enzyme specificity was set, peptide mass error tolerances were set at 5 ppm for precursors and 0.03 Da for MS2 fragments. Additionally, post-translational modifications were identified utilizing the PEAKS inbuilt *de novo*-led search for 303 common modifications. FDR was calculated using the decoy database search built into PEAKS (Table S11).

## Supplemental information

### JDRF Diabetes, Genes, Autoimmunity, and Prevention (D-GAP) cohort

**Table S1:** D-GAP sample selection summary, with samples grouped by HLA DR-DQ diplotypes and sorted by T1D risk, measured as the log_10_-transformed odds ratio (OR) of developing T1D contributed by each diplotype. OR estimates were obtained from UK Biobank, ^24^ and used as the explanatory variable (Generative model 1). The categorical group variable only has a nominal value. AA_1_ and AA_2_ encode amino acids at HLA DQβ1_57_, DRβ1_13_, and DRβ1_71_. ^6^

| n | Haplotype 1 |  |  | Haplotype 2 |  |  | AA<sub>1</sub> | AA<sub>2</sub> | log(OR) | Group |
| --- | --- | --- | --- | --- | --- | --- | --- | --- | --- | --- |
|  | DRB1 | DQA1 | DQB1 | DRB1 | DQA1 | DQB1 |  |  |  |  |
| 2 | 15:01 | 01:02 | 06:02 | 11:04 | 05:05 | 06:03 | DRA | DSR | -1.30 | Protected |
| 1 | 15:01 | 01:02 | 06:02 | 13:02 | 01:02 | 06:04 | DRA | VSE | -1.30 | Protected |
| 2 | 15:01 | 01:02 | 06:02 | 16:01 | 01:02 | 05:02 | DRA | SRR | -1.30 | Protected |
| 2 | 15:01 | 01:02 | 06:02 | 15:01 | 01:02 | 06:02 | DRA | DRA | -0.96 | Protected |
| 1 | 15:01 | 01:02 | 06:02 | 11:04 | 05:05 | 03:01 | DRA | DSR | -0.89 | Protected |
| 1 | 15:01 | 01:02 | 06:02 | 12:01 | 05:05 | 03:01 | DRA | DGR | -0.89 | Protected |
| 1 | 04:01 | 03:01 | 03:01 | 11:03 | 05:05 | 03:01 | DHK | DSE | 0.09 | Low risk |
| 1 | 04:07 | 03:03 | 03:01 | 01:03 | 05:05 | 03:01 | DHR | DFE | 0.09 | Low risk |
| 3 | 04:01 | 03:02 | 03:01 | 04:01 | 03:02 | 03:01 | DHK | DHK | 0.58 | Low risk |
| 1 | 04:08 | 03:02 | 03:01 | 04:07 | 03:02 | 03:01 | DHR | DHR | 0.58 | Low risk |
| 1 | 04:01 | 03:01 | 03:01 | 12:02 | 06:01 | 03:01 | DHK | DGR | 0.59 | Low risk |
| 1 | 09:01 | 03:02 | 02:02 | 04:01 | 03:02 | 03:01 | AFR | DHK | 0.59 | Low risk |
| 5 | 04:01 | 03:01 | 03:02 | 04:01 | 03:01 | 03:02 | AHK | AHK | 1.51 | Susceptible |
| 1 | 04:04 | 03:01 | 03:02 | 04:01 | 03:01 | 03:02 | AHR | AHK | 1.51 | Susceptible |
| 2 | 04:04 | 03:01 | 03:02 | 04:04 | 03:01 | 03:02 | AHR | AHR | 1.51 | Susceptible |
| 17 | 04:01 | 03:01 | 03:02 | 03:01 | 05:01 | 02:01 | AHK | ASK | 1.80 | Susceptible |
| 1 | 04:02 | 03:01 | 03:02 | 03:01 | 05:01 | 02:01 | AHE | ASK | 1.80 | Susceptible |
| 1 | 04:03 | 03:01 | 03:02 | 03:01 | 05:01 | 02:01 | AHR | ASK | 1.80 | Susceptible |
| 4 | 04:04 | 03:01 | 03:02 | 03:01 | 05:01 | 02:01 | AHR | ASK | 1.80 | Susceptible |

**Table S2:**
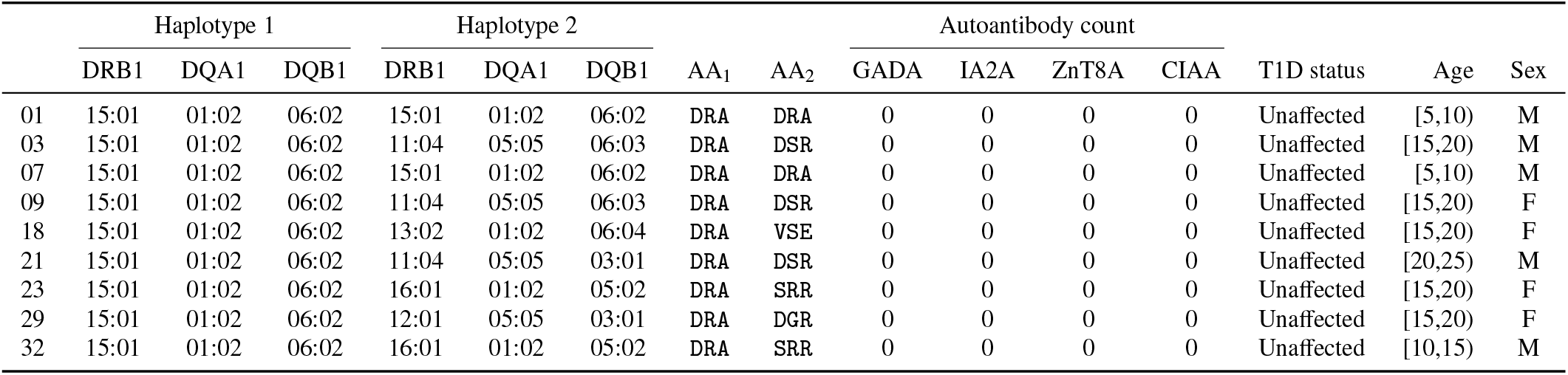
D-GAP genetically protected group HLA DR-DQ diplotypes, autoantibody count, and disease status at the time of peripheral blood sample collection. AA_1_ and AA_2_ encode amino acids at HLA DQβ1_57_, DRβ1_13_, and DRβ1_71_. ^6^

**Table S3:** D-GAP low genetic risk group HLA DR-DQ diplotypes, autoantibody count, and disease status at the time of peripheral blood sample collection. AA_1_ and AA_2_ encode amino acids at HLA DQβ1_57_, DRβ1_13_, and DRβ1_71_. ^6^ The DQA1*03:02 allele is unresolved to DQA1*03:02 or DQA1*03:03 due to insufficient HLA typing resolution.

|  | Haplotype 1 |  |  | Haplotype 2 |  |  | AA<sub>1</sub> | AA<sub>2</sub> | Autoantibody count |  |  |  | T1D status | Age | Sex |
| --- | --- | --- | --- | --- | --- | --- | --- | --- | --- | --- | --- | --- | --- | --- | --- |
|  | DRB1 | DQA1 | DQB1 | DRB1 | DQA1 | DQB1 |  |  | GADA | IA2A | ZnT8A | CIAA |  |  |  |
| 42 | 09:01 | 03:02 | 02:02 | 04:01 | 03:02 | 03:01 | AFR | DHK | 0 | 0 | 0 | 0 | Unaffected | [5,10) | F |
| 43 | 04:07 | 03:03 | 03:01 | 01:03 | 05:05 | 03:01 | DHR | DFE | 0 | 0 | 0 | 0 | Unaffected | [10,15) | M |
| 45 | 04:01 | 03:02 | 03:01 | 04:01 | 03:02 | 03:01 | DHK | DHK | 0 | 0 | 0 | 0 | Unaffected | [10,15) | M |
| 46 | 04:01 | 03:01 | 03:01 | 12:02 | 06:01 | 03:01 | DHK | DGR | 0 | 0 | 0 | 0 | Unaffected | [15,20) | F |
| 51 | 04:01 | 03:02 | 03:01 | 04:01 | 03:02 | 03:01 | DHK | DHK | 0 | 0 | 0 | 0 | Unaffected | [5,10) | M |
| 52 | 04:08 | 03:02 | 03:01 | 04:07 | 03:02 | 03:01 | DHR | DHR | 0 | 0 | 0 | 0 | Unaffected | [5,10) | M |
| 53 | 04:01 | 03:01 | 03:01 | 11:03 | 05:05 | 03:01 | DHK | DSE | 0 | 0 | 0 | 0 | Unaffected | [20,25) | M |
| 55 | 04:01 | 03:02 | 03:01 | 04:01 | 03:02 | 03:01 | DHK | DHK | 2 | 0 | 2 | 2 | Unaffected | [5,10) | M |

**Table S4:**
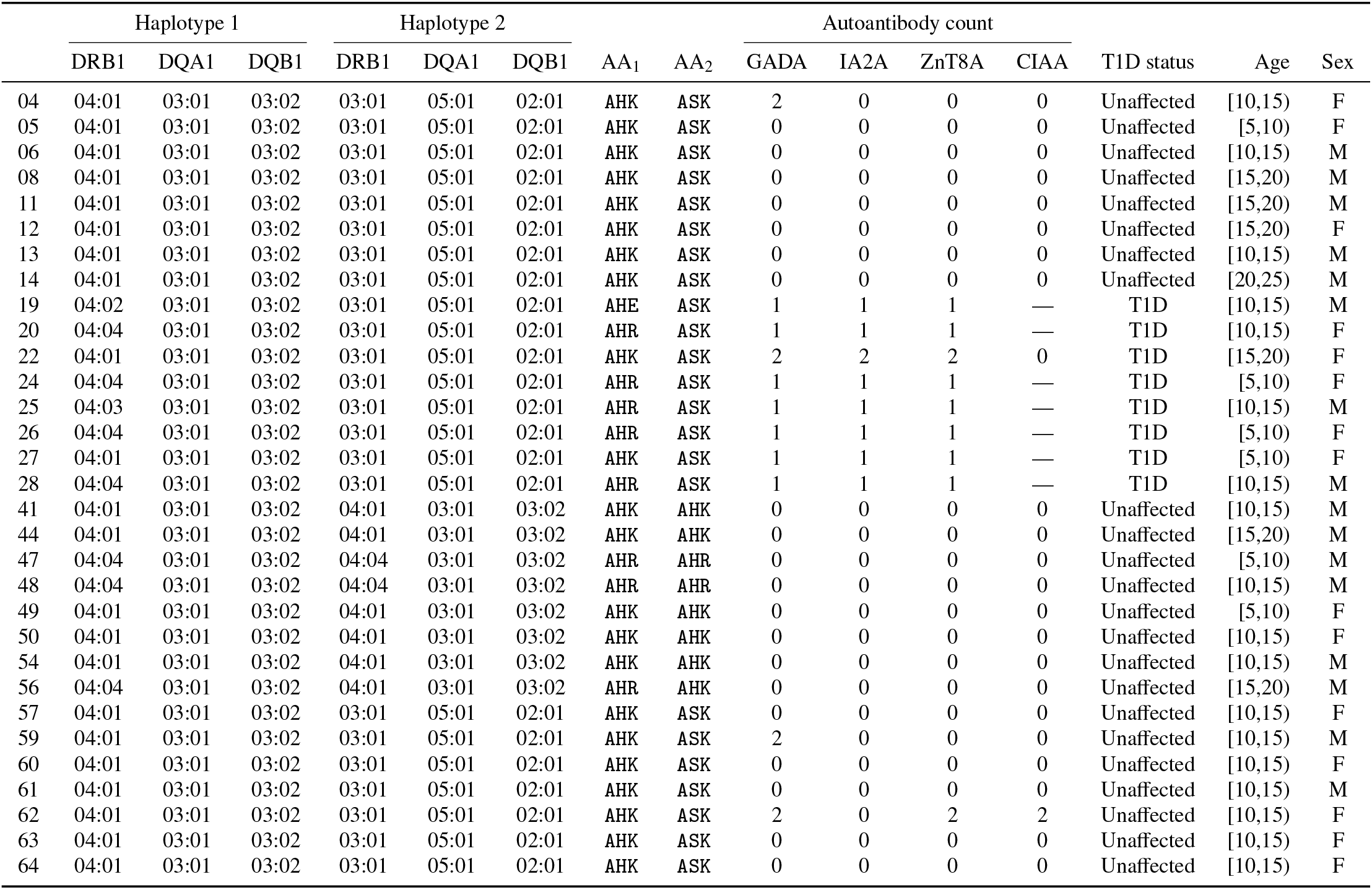
D-GAP genetically susceptible group HLA DR-DQ diplotypes, autoantibody count, and disease status at the time of peripheral blood sample collection. AA_1_ and AA_2_ encode amino acids at HLA DQβ1_57_, DRβ1_13_, and DRβ1_71_. ^6^ — denotes missing data.

**Table S5:** Top 5 gene markers for single CD4^+^ T cell clusters 0–11 in the D-GAP cohort (Figure S1). Genes labeled with ^*^ indicate no standard name is currently assigned, and the identifier of the corresponding genomic region has been used as a placeholder.

|  | Gene marker rank |  |  |  |  |
| --- | --- | --- | --- | --- | --- |
|  | 1 | 2 | 3 | 4 | 5 |
| 0 | CD40LG | NFKBID | SLA | H3F3B | PRNP |
| 1 | AC083862* | AL590652* | AP001160* | HSD17B1 | LINC00926 |
| 2 | FOS | DNA1B1 | TSC22D3 | JUN | DUSP1 |
| 3 | TXNIP | CXCL3 | TMSB10 | CCL4 | SH2D1B |
| 4 | IL13 | IFNG | TNF | CCL5 | IL2 |
| 5 | C17orf100 | CYTOR | CNTLN | AG01000058* | CLDND1 |
| 6 | RPS2 | RPS27 | RGCC | PIM3 | DUSP2 |
| 7 | CXCL3 | CXCL8 | CXCL2 | CLDN1 | CCR4 |
| 8 | FOXP3 | CTLA4 | TNFRSF9 | IKZF2 | TIGIT |
| 9 | MALAT1 | SRGN | CD69 | IL7R | HLA-B |
| 10 | CRTAM | XCL2 | XCL1 | KLRK1 | CSF2 |
| 11 | CLPTM1L | CRBN | ZNF519 | AKNA | SAMD10 |

**Figure S1:**
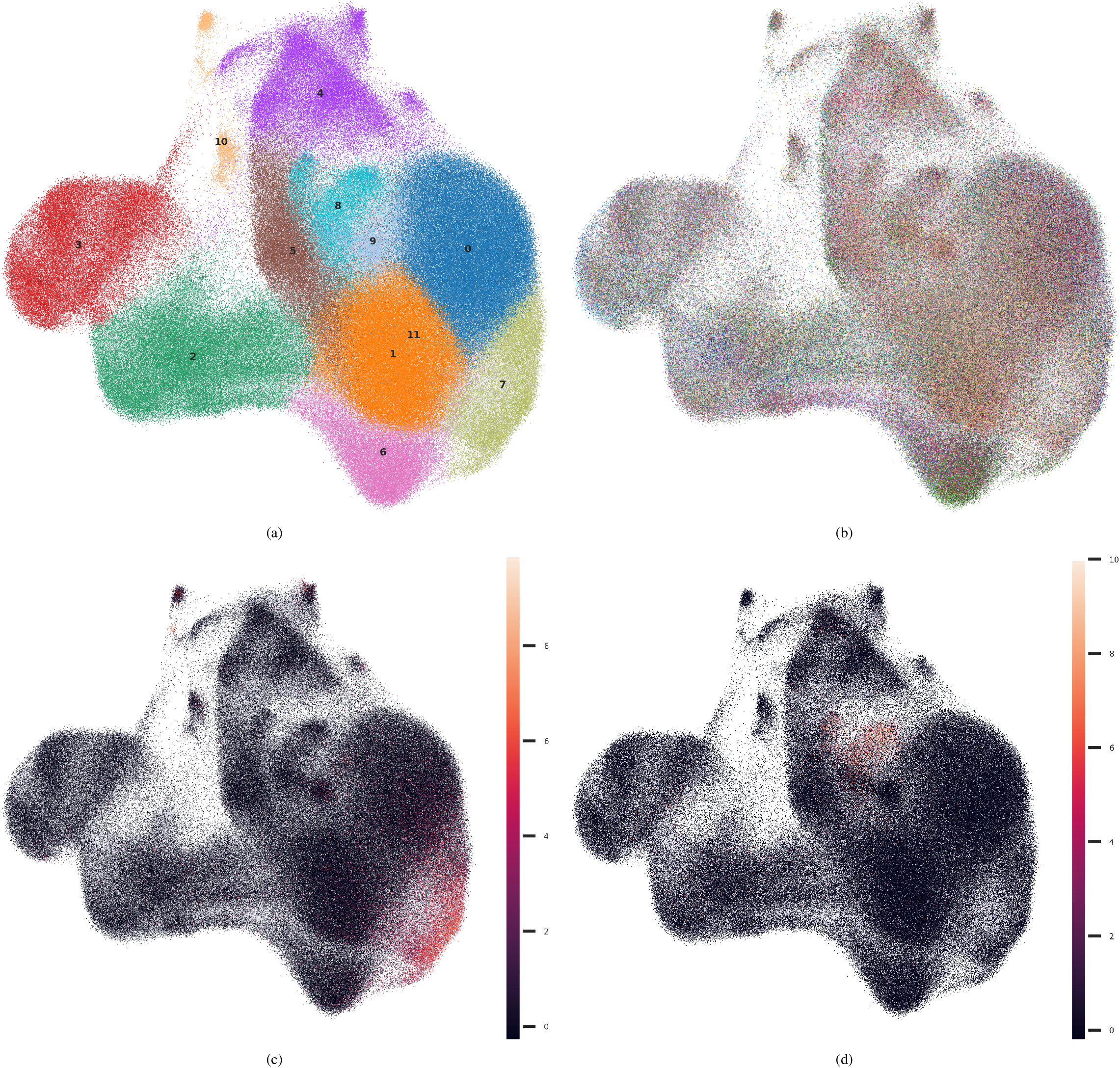
Single-cell RNA subsets in the D-GAP cohort. Each point represents one cell. Expression levels are indicated in log_10_-transformed read counts, where 1 unit = 10^4^ reads prior to the transformation. (a) CD4^+^ subsets 0–11. (b) Donor distribution. (c) *CXCL8* expression, a marker of recent thymic emigrants (RTEs), ^25^ identified as subset 7. (d) *FOXP3* expression, a marker of regulatory T cells (Tregs), identified as subset 8. CD4^+^ T conventional cells (Tconvs) were defined as the union of all subsets except 7 and 8.

**Figure S2:**
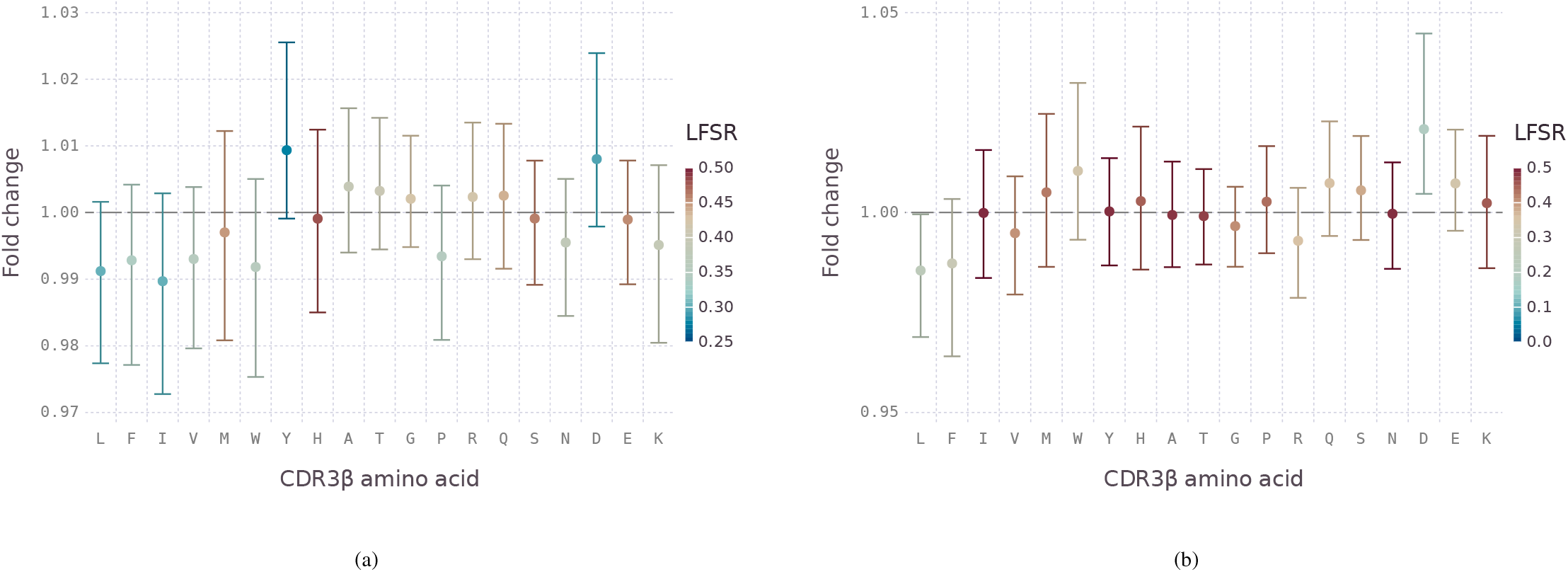
Estimates of T1D odds ratio effects, contributed by HLA DR-DQ diplotypes, on CDR3β 1-mers from recent thymic emigrant (RTE) and regulatory (Treg) CD4^+^ T cells, excluding cysteine. Error bars summarize the effects on multiple k-mers by depicting the median and 90% credible intervals of fold changes across HLA class II extremes, with the most protective DQ6 diplotypes as baseline (Table S1). (a) RTE cell subset. (b) Treg cell subset.

**Figure S3:**
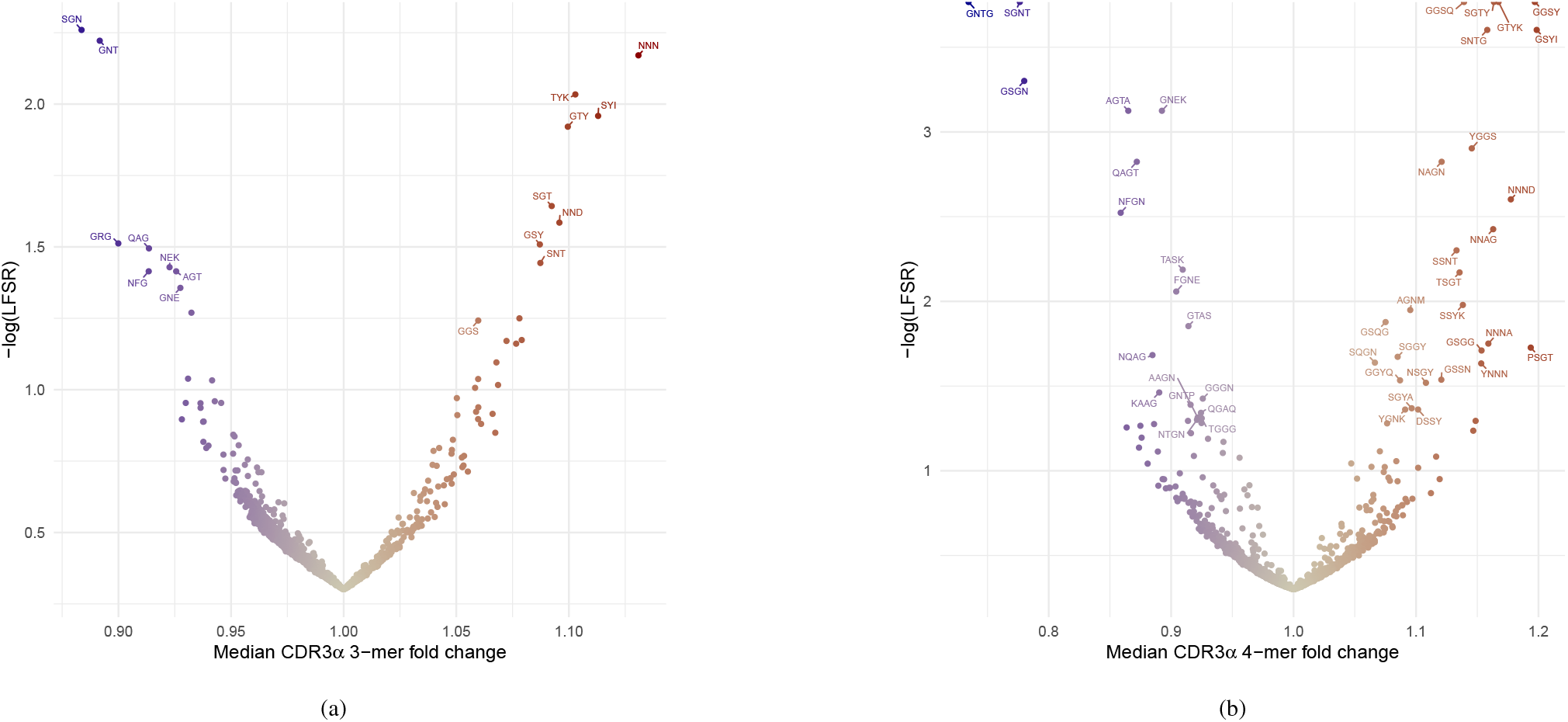
Estimates of T1D odds ratio effects, contributed by HLA DR-DQ diplotypes, on CDR3α k-mers from CD4^+^ T conventional cells. Funnel plots represent the median and the local false sign rate (LFSR) of fold changes across HLA class II extremes, with the most protective DQ6 diplotypes as baseline (Table S1). (a) All 3-mers with non-zero counts. (b) All 4-mers with non-zero counts. 4-mers on the y axis boundary have an estimated LFSR ≈ 0.

### Investigating Underlying Causal Mechanisms of Type 1 Diabetes (DMech) donors

**Table S6:** DMech donor HLA-DR-DQ diplotypes and autoantibody status at the time of peripheral blood sample collection.

|  | Haplotype 1 |  |  | Haplotype 2 |  |  | IAA |  | GADA |  | ICA | Age | Sex |
| --- | --- | --- | --- | --- | --- | --- | --- | --- | --- | --- | --- | --- | --- |
|  | DRB1 | DQA1 | DQB1 | DRB1 | DQA1 | DQB1 | Status | U/ml | Status | U/ml |  |  |  |
| 11 | 04:01 | 03:01 | 03:02 | 04:02 | 03:01 | 03:02 | Negative | 0.80 | Positive | 15.9 | Positive | [5,10) | M |
| 13 | 03:01 | 05:01 | 02:01 | 03:01 | 05:05 | 02:02 | Negative | 0.80 | Positive | 91.9 | Positive | [10,15) | M |
| 14 | 04:01 | 03:01 | 03:02 | 04:01 | 03:01 | 03:02 | Positive | 6.34 | Positive | 86.3 | Positive | [5,10) | F |
| 16 | 03:01 | 05:01 | 02:01 | 03:01 | 05:05 | 02:02 | Positive | 8.44 | Positive | 88.2 | Positive | [10,15) | M |
| 18 | 04:01 | 02:01 | 02:02 | 07:01 | 03:01 | 03:02 | Positive | 6.14 | Positive | 71.8 | Positive | [5,10) | F |

### Network of Pancreatic Organ Donors with Diabetes (nPOD)

**Table S7:** nPOD donor HLA DR-DQ diplotypes, autoantibody status, number of preproinsulin-reactive (PPI) CD4^+^ T cell clones isolated from the pancreas, and years since diagnosis at the time of death. — denotes missing data.

|  | Haplotype 1 |  |  | Haplotype 2 |  |  | Autoantibody |  |  |  | PPI-reactive |  | Diag. | Age | Sex |
| --- | --- | --- | --- | --- | --- | --- | --- | --- | --- | --- | --- | --- | --- | --- | --- |
|  | DRB1 | DQA1 | DQB1 | DRB1 | DQA1 | DQB1 | IAA | IA2A | GADA | ZnT8A | Positive | Tested |  |  |  |
| 69 | 04:01 | 03:01 | 03:02 | 07:01 | 02:01 | 02:02 | — | — | — | — | 0 | 7 | [0,5) | [5,10) | F |
| 6323 | 03:01 | 05:01 | 02:01 | 04:02 | 03:01 | 03:02 | Negative | Positive | Positive | Negative | 4 | 52 | [5,10) | [20,25) | F |
| 6342 | 01:01 | 01:01 | 05:01 | 04:01 | 03:01 | 03:02 | Positive | Positive | Negative | Negative | 5 | 34 | [0,5) | [10,15) | F |
| 6367 | 04:01 | 03:01 | 03:02 | 07:01 | 02:01 | 02:02 | Negative | Negative | Negative | Negative | 0 | 9 | [0,5) | [20,25) | M |
| 6414 | 03:01 | 05:01 | 02:01 | 09:01 | 03:03 | 02:02 | Positive | Negative | Positive | Positive | 4 | 34 | [0,5) | [20,25) | M |
| 6472 | 03:01 | 05:01 | 02:01 | 04:04 | 03:01 | 03:02 | Positive | Negative | Negative | Negative | 1 | 30 | [0,5) | [10,15) | F |
| 6533 | 03:01 | 05:01 | 02:01 | 04:01 | 03:01 | 03:02 | Positive | Positive | Negative | Positive | — | — | [0,5) | [0,5) | F |
| 6536 | DR4 | DQ8 | DQ8 | DR17 | DQ2 | DQ2 | Negative | Negative | Positive | Negative | — | — | [0,5) | [20,25) | F |

### Insulin mimotope discovery

**Table S8:**
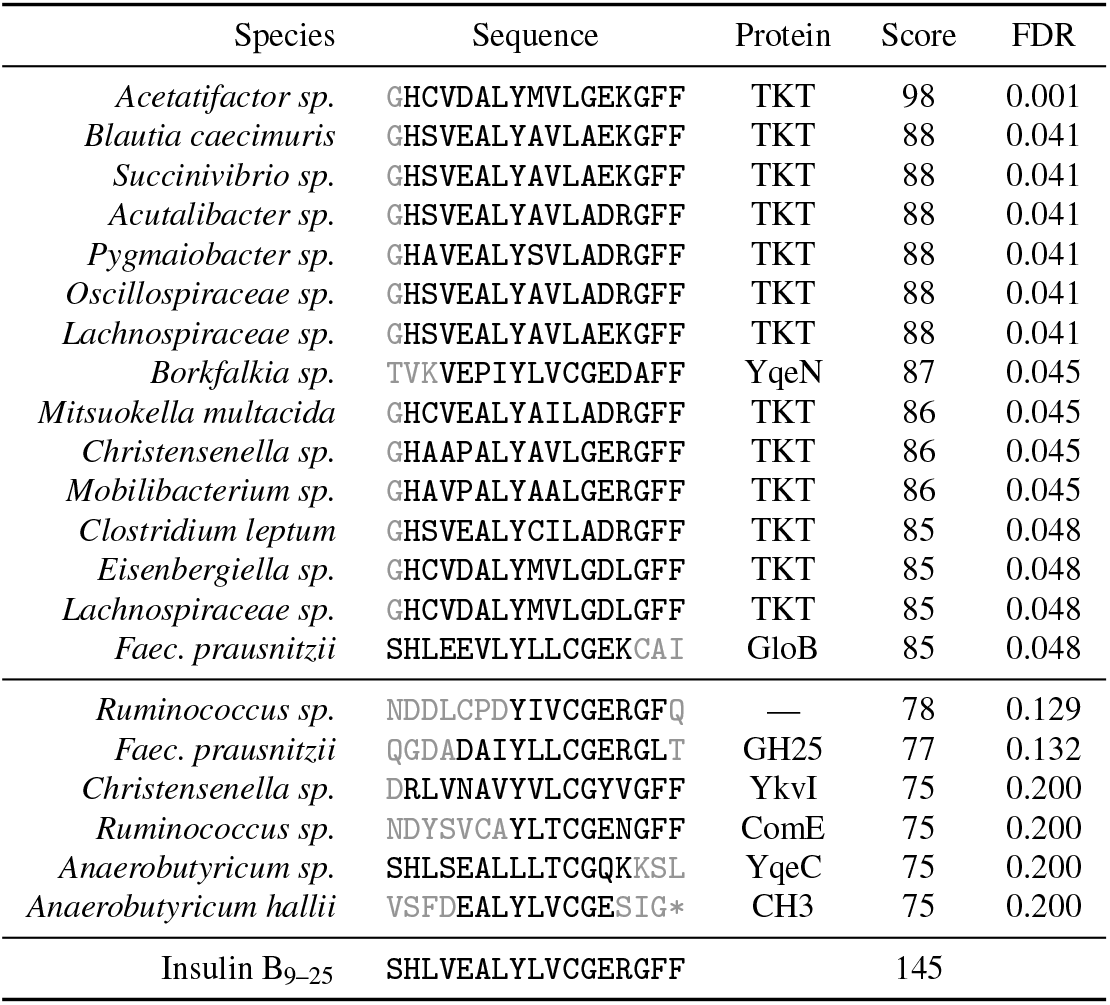
Top scoring local alignments to insulin B_9–25_ in proteins from the gut microbiome, FDR ≤ 0.05. Top scoring members from other protein families, FDR ≤ 0.2, are also included. Shaded residues do not belong to the optimal local alignment to insulin B_9–25_. Protein domains and superfamilies listed include N-terminal transketolase (TKT), yqeN DNA replication protein (YqeN), hydroxyacylglutathione hydrolase (GloB), glycosyl hydrolase family 25 (GH25), uncharacterized membrane protein (YkvI), late competence operon (ComE), selenium cofactor biosynthesis protein (YqeC), and cyclases/histidine kinases associated sensory extracellular domain 3 (CH3). — indicates no protein family could be predicted. For reference, human insulin paralogs insulin-like growth factor I (IGF-1) and II (IGF-2) scores are 90 and 81, respectively, whereas human transketolase has a score of 43.

**Figure S4:**
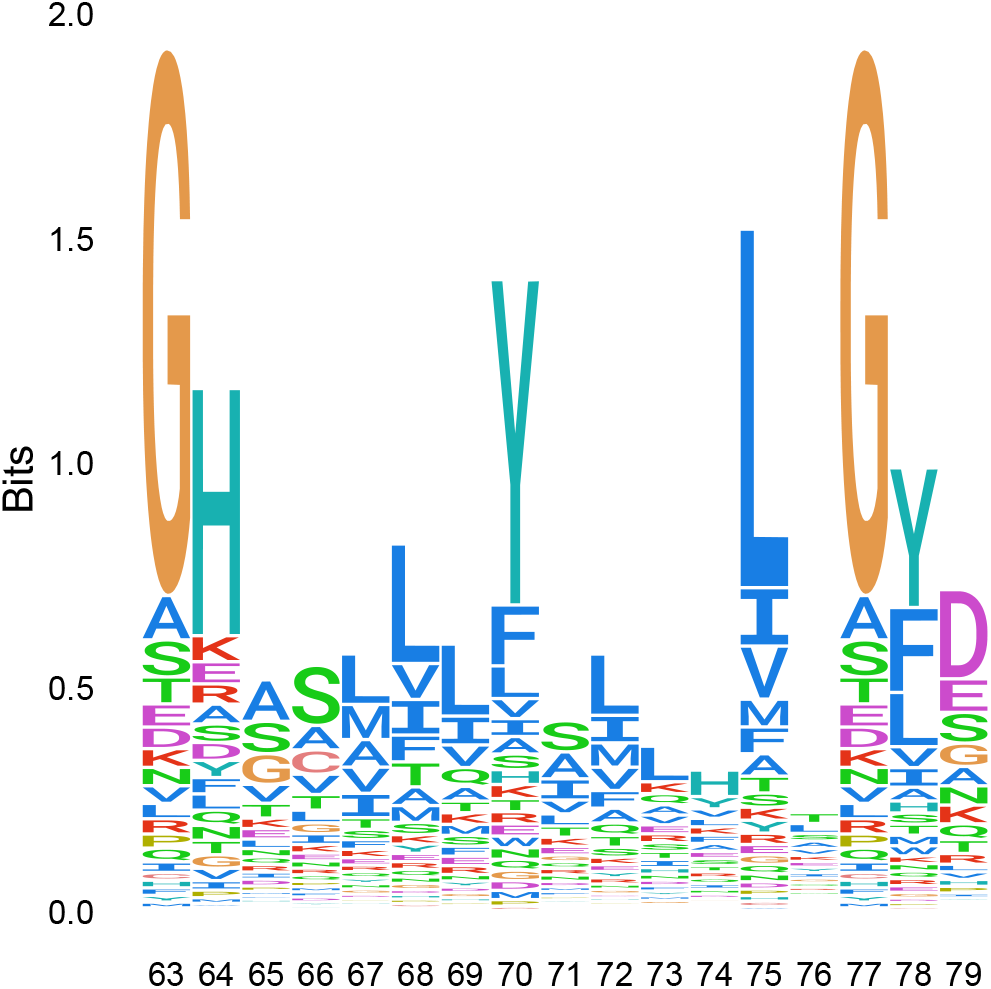
N-terminal transketolase domain (Pfam PF00456) motif for all domains of life in the region of insulin mimicry. A large information content in bits denotes strong conservation. Most conserved residues in transketolase (TKT) are similar or identical to those from the corresponding insulin B_9–25_ (SHLVEALYLVCGERGFF) position. This may facilitate the evolution of insulin B_9–25_ mimotopes using TKT as a template.

### CD4^+^ T cell stimulation assays

**Table S9:**
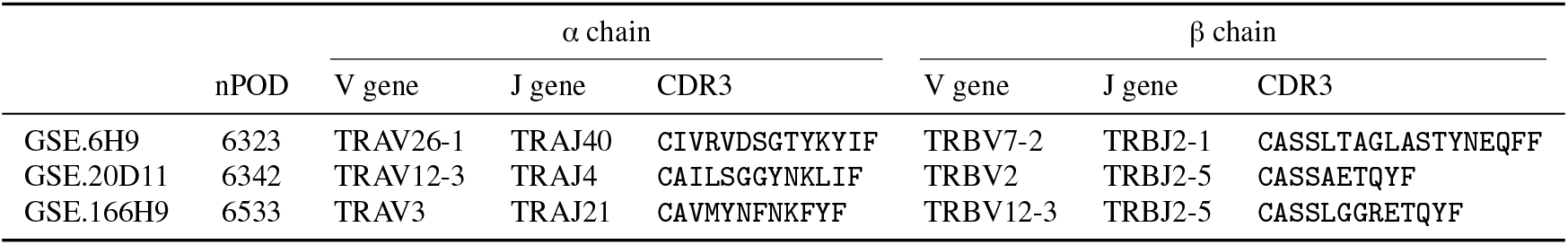
nPOD CD4^+^ T cell clonotypes stimulated by insulin B_9–23_ (Figure 4). GSE.6H9 and 20D11 were previously reported. ^33^ GSE.166H9 is a novel clonotype that cross-reacts with transketolase from *Clostridium leptum*, peptide 8 (Table S10).

**Table S10:**
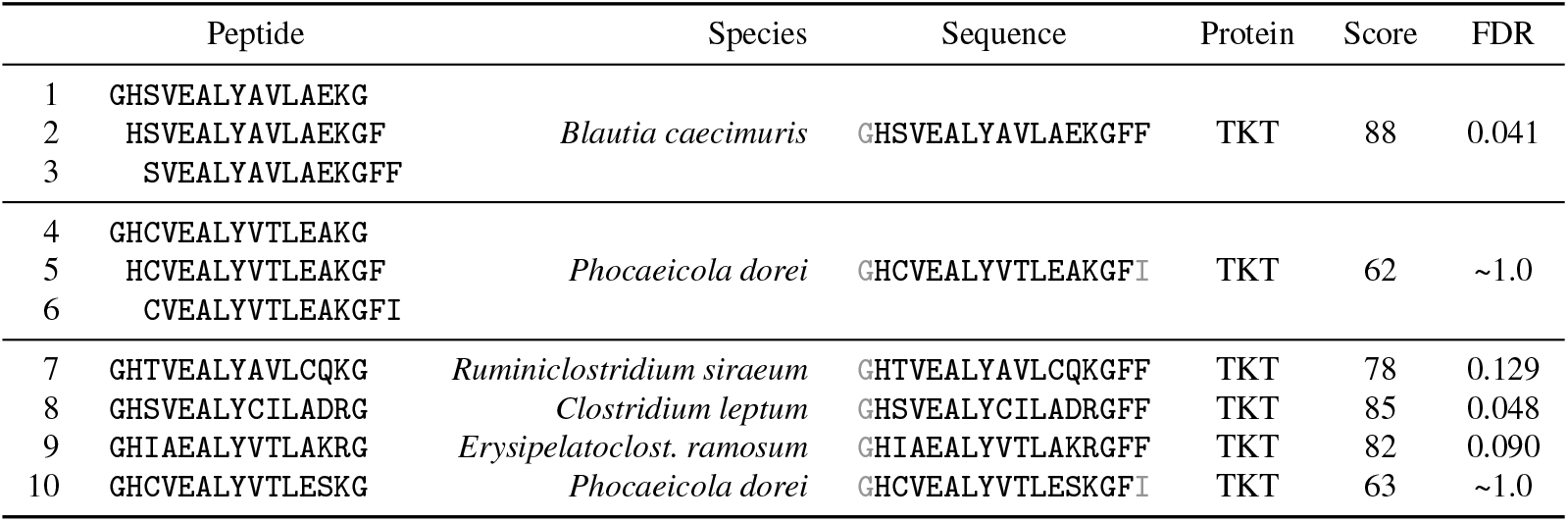
Peptides employed on CD4^+^ T cell stimulation assay (Figure 4), derived from prioritized mimotopes (Table S8). Shaded residues do not belong to the optimal local alignment to insulin B_9–25_. Peptide 8 stimulates the novel anti-insulin clonotype GSE.166H9 (Table S9). Peptides were chosen to maximize sequence diversity within transketolase sequences with FDR ≤ 0.2 and also present in >10% individuals from IGC, a population-wide gut microbiome reference catalog. ^67^ Despite lower similarity to insulin B_9–25_, *P. dorei* peptides were also included owing to the increase in abundance of this species observed prior to autoimmunity in one cohort study. ^18^

### Peptide elution in HLA-DQ molecules

**Table S11:** Eluted peptides from *Blautia caecimuris* transketolase measured to be bound by HLA-DQ molecules using mass spectrometry, FDR ≤ 0.05. Using an anti-DQ antibody, peptides presented by EBV B-cell lines were identified from elution experiments with either DQ6^+^ or DQ8^+^ cells. Peptides FVMSKGHSVEALY and GHSVEALYAVLAE, bound by DQ6, correspond to the region of insulin B_9–25_ mimicry (Figure S5).

| Sample | Peptide | *P* | FDR |
| --- | --- | --- | --- |
| DQ6 | GKPTVLIANTVKGCGSSVMENK | $1.74 \times 10^{-6}$ | 0.029 |
| DQ6 | KDVINMIRSGKAGHIG | $1.20 \times 10^{-5}$ | 0.029 |
| DQ6 | FVMSKGHSVEALY | $1.90 \times 10^{-5}$ | 0.029 |
| DQ6 | IEEVIDTFSKFGSK | $5.26 \times 10^{-5}$ | 0.029 |
| DQ6 | GHSVEALYAVLAE | $2.92 \times 10^{-4}$ | 0.029 |
| DQ6 | CVGMALAGK | $1.85 \times 10^{-3}$ | 0.029 |
| DQ8 | VTLYFKQMNISPEN | $1.01 \times 10^{-5}$ | 0.025 |
| DQ8 | LDAAFEEAKTVKGKPTVL | $4.35 \times 10^{-5}$ | 0.025 |
| DQ8 | NSIEELDAAFEEAKT | $1.04 \times 10^{-3}$ | 0.025 |

**Figure S5:**
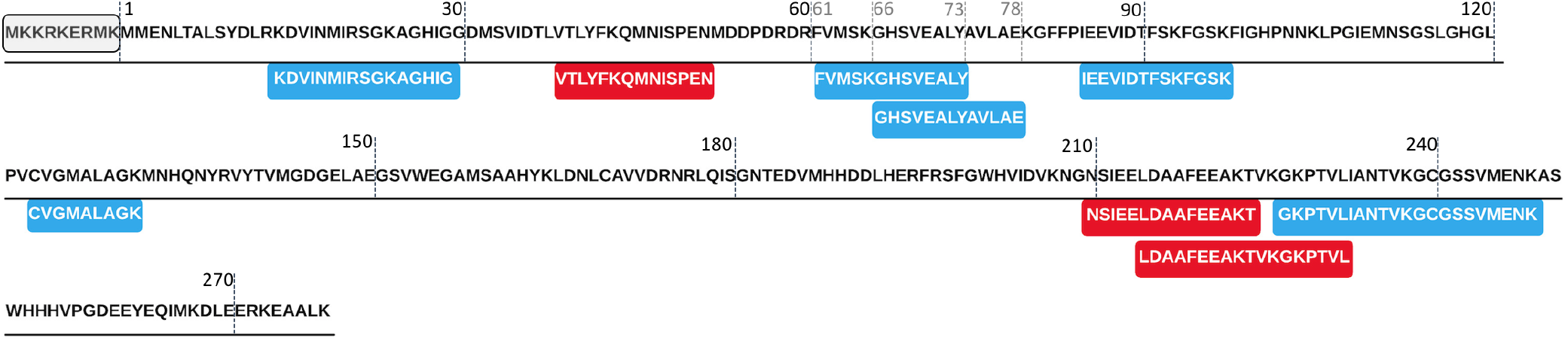
Eluted peptides measured to be bound by HLA-DQ6 (blue) and DQ8 (red) molecules using mass spectrometry, FDR ≤ 0.05 (Table S11), overlaid on the *Blautia caecimuris* transketolase sequence they originate from. Peptides in the region 61–78 correspond to insulin B_9–25_ mimics. The N-terminal protein sequence (`MKKRKERMK`), present in the IGC gut metagenome assembly, ^67^ is absent in the alternative assembly constructed by MGnify. ^31^ The longer version was synthesized and used in the elution assays.

